# Language-network connectivity and cortical glutamate and their associations with overweighted semantic prior beliefs, schizotypy and task-based hallucinations

**DOI:** 10.64898/2025.12.07.25341628

**Authors:** Verena F. Demler, Elisabeth F. Sterner, Franziska Knolle

## Abstract

**Background:** Impairments in language comprehension and production are core symptoms of schizophrenia spectrum disorders (SSD). Predictive processing, a Bayesian framework of brain function, provides an explanatory account of both language processing and psychotic symptom formation. Within SSD, neurobiological correlates of alterations in predictive language processing are poorly understood. Here, we investigate if glutamatergic neurotransmission and functional connectivity (FC) within the language network are associated with (1) altered predictive language processing, (2) task-based hallucinations (TBH), and (3) schizotypy.

**Methods:** In 53 healthy individuals (26 women), we combined a predictive language task analyzed using a Bayesian belief update model to estimate the individuals’ reliance on prior knowledge versus sensory input depending on the their schizotypal trait expression, resting-state functional MRI, and MR spectroscopy to measure glutamate levels in the anterior cingulate cortex (ACC) and left dorsolateral prefrontal cortex (DLPFC). Seed-based FC analysis focused on the language network including the left inferior frontal gyrus (IFG), posterior superior temporal gyrus (pSTG), and posterior middle temporal gyrus (pMTG).

**Results:** Increased FC between left IFG and frontal areas predicted schizotypy-informed prior overweighting. Enhanced left pSTG-medial frontal FC was associated with the frequency of TBH. Finally, schizotypy was predicted by interactions between ACC/DLPFC Glx and left pSTG-frontal FC.

**Conclusions:** These results provide novel insights into neurobiological and computational mechanisms underlying altered language-related predictive processing and subclinical symptom formation. They highlight potential early indicators of schizotypal symptom development and underscore the value of multimodal approaches in elucidating pathophysiology of SSD.

## 1 Introduction

Impairments in language comprehension and production are core features of schizophrenia spectrum disorders (SSDs) and recent studies indicated that these may also contribute to the development of psychotic symptoms(1–4). However, the underlying neurobiological and computational mechanisms remain poorly understood.

From a computational perspective, both language processing deficits and psychotic symptoms can be conceptualized within the framework of predictive processing. This theory proposes that cognitive processes, including language(4–8), may rely on probabilistic inference. At each cortical level, predictions, based on prior beliefs, are compared with incoming sensory evidence at the lowest level. The prediction error is then propagated upwards to update beliefs and minimize future discrepancies(9–11). This process involves weighing prior beliefs and sensory information according to their precision (reliability) to optimize the inference under uncertainty. Imbalances in the weighting process may distort language perception, potentially leading to misperceptions or hallucinations. Consistent with this, predictive processing models of psychosis suggest that such imbalances may underlie the development of psychotic symptoms (e.g., hallucinations and delusions)(12–15). Yet, findings remain inconsistent, with studies reporting both, an overweighting of prior beliefs(e.g., (16–18)) or of sensory evidence(e.g., (19–22)).

In a previous study(18), we tackled these inconsistencies by developing a predictive language task that simultaneously manipulated the precision of prior beliefs and sensory evidence. By applying a mechanistic Bayesian belief updating model, we found that individuals with increasing schizotypal traits overweighed prior beliefs and experienced more task-based hallucinations (TBH). On a neurobiological level, increased ACC glutamate significantly predicted the increasing prior weight(18), consistent with theories that link glutamatergic dysregulation to altered weighting of priors(10,15,23). We focused on schizotypy, as schizophrenia is increasingly viewed as a continuum, ranging from subclinical expressions to severe clinical manifestations(24,25). Schizotypy not only shares similar biological alterations with schizophrenia, though more subtle(25,26), but also displays similar cognitive alterations, specifically in language processing(27–29). As a potential risk marker for developing SSDs, schizotypy allows investigating potential neurobiological alterations within subclinical stages and could provide a better understanding of the disorder’s clinical onset(30). However, a comprehensive understanding of how clinical symptoms arise from pathophysiological alterations requires a combination of different neurobiological approaches.

The present study, therefore, aimed to investigate whether cortical glutamatergic neurotransmission interacts with resting-state FC within the language network to predict prior overweighting, as a computational marker, during language processing. Additionally, we explored whether potential neurobiological alterations related to a phenomenological marker, the TBH frequency, and to schizotypy, as a trait marker of schizophrenia. Resting-state FC is particularly relevant given the extensive evidence characterizing schizophrenia as a disorder of functional dysconnectivity marked by hypo- and hyper-functional integration between brain regions(31–33). Critically, these alterations are evident in early disease stages(34–36) and even precede clinical onset(37–39). Notably, frontotemporal brain regions, including the inferior frontal gyrus (IFG), the superior (STG) and middle temporal gyri (MTG), and the inferior parietal lobule, exhibit hyperactivity and altered connectivity(34,38,40,41), which may predict psychosis onset(42–44). Furthermore they are integral to the language network, underlying language production and comprehension(4,45,46) and have been shown to be involved in the development of auditory-verbal hallucinations, underscoring the link between language and psychotic symptoms(47–49). Given the left language lateralization in most right-handed subjects(45,50), we focused on the left IFG, left posterior STG (pSTG), and left posterior MTG (pMTG).

Previous research furthermore proposed altered glutamate signaling as a pathophysiological mechanism underlying abnormal FC, due to its critical role in regulating excitation-inhibition(E/I)-balance, which in turn modulates synchronized slow fluctuations of neural activity that can be captured in resting-state fMRI(35,36,51). Importantly, glutamate dysregulations have been reported across different disease stages(52–55).

Integrating these findings, this study investigates the interplay between resting-state FC in the language network and cortical glutamate levels in predicting imbalances in prior weighting during language perception, the occurrence of TBH, and schizotypal traits. By combining computational, neurobiological, and phenomenological approaches, we aimed to elucidate the mechanisms underlying subclinical symptom development in schizotypy.

## 2 Methods and Materials

### 2.1 Participants

In this study we included 53 healthy subjects (26 women) aged between 18-35 years (mean age=23.62±3.87). All participants completed a German Likert-version(56) of the Schizotypal Personality Questionnaire (SPQ)(57) capturing schizotypy (SPQ scores range from 16-209, mean SPQ score=88.47 ±49.85). For further demographic and clinical information, recruitment and inclusion criteria refer to the supplementary material and previous papers(18,52,58). In this study, participants first completed a predictive language task behaviorally on a laptop. Subsequently, they underwent MRI scanning, including structural, functional, and spectroscopy scans. The study was approved by the medical research ethics committee of the Technical University of Munich. All subjects gave written informed consent in accordance with the Declaration of Helsinki.

### 2.2 Predictive language task

During the predictive language task, participants first listened to a sentence and rated the clarity of a degraded sentence’s final word (i.e., target word). Then they reported the target word and rated their confidence in their answer. In total, each participant completed 200 sentence trials. Sentence stimuli were selected from the MuSe database(59). We modulated the predictability (i.e., entropy, low entropy means thereby highly predictable and the other way round) of the sentence, as well as the surprisal (i.e., cloze probability) and the level of sensory detail (i.e., noise degradation, four levels) of the target word. The task parameters were implemented in a computational belief updating model described in section 2.6.

### 2.3 MR Image acquisition

All participants underwent a structural MRI and ^1^H-MRS scan on a 3T Philips Ingenia Elition X MR-Scanner (Philips Healthcare, Best, The Netherlands) using a 32-channel head coil. For anatomical reference and spectroscopic voxel placement, we acquired a T1-weighted magnetization prepared rapid gradient echo (MPRAGE) sequence. MRS data were collected using a ¹H-MRS single voxel ECHO volume Point Resolved Spectroscopy Sequence (PRESS) sequence. The voxels were placed in the ACC (20x20x20mm) and left DLPFC (30x20x20mm); see Figure 1 for the overlap in voxel placement. Resting-state fMRI was acquired during the same session with a multiband-SENSE sequence. For further descriptions see supplementary material and previous papers(52,58).

**Figure 1:**
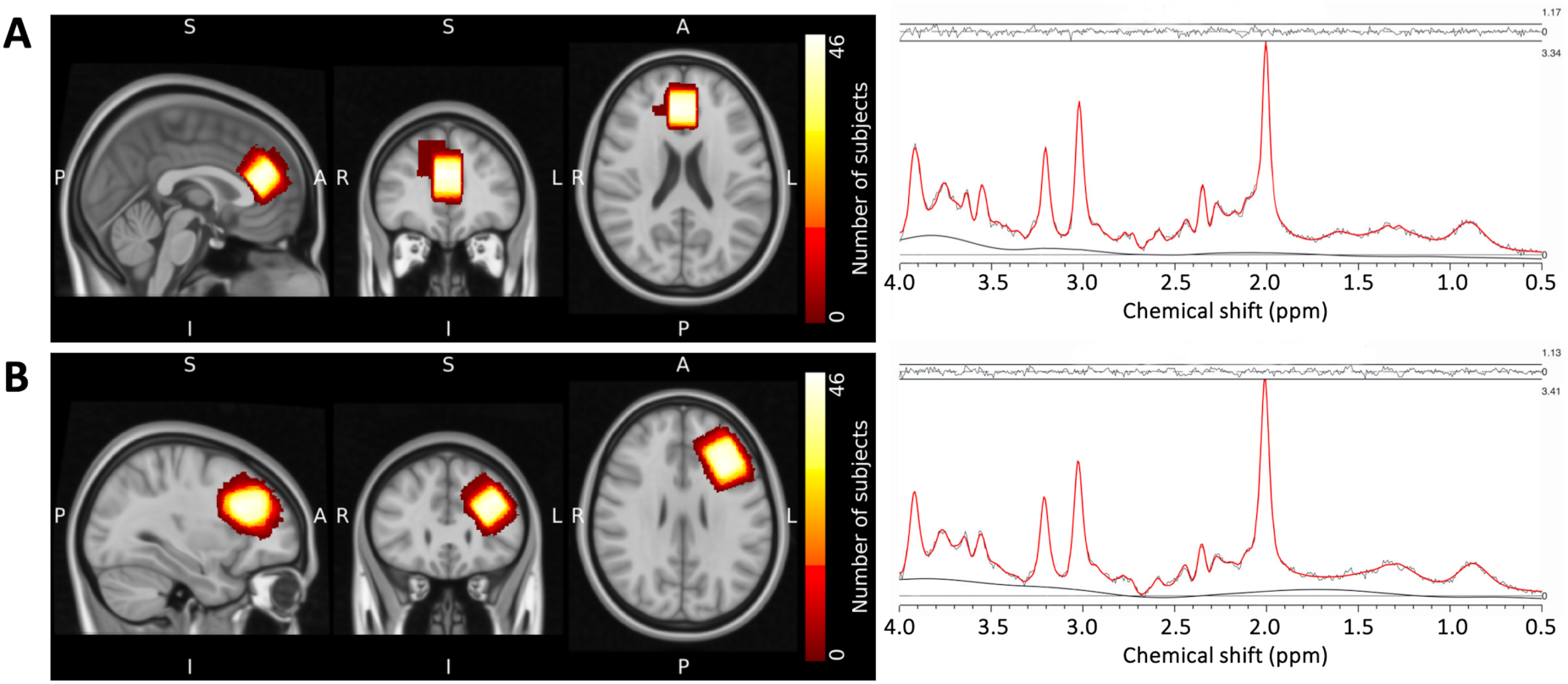
Voxel placement and representative fitted MRS spectra. Note: Placement of MRS voxels and 1H-MRS spectrum fitted by Osprey of the (A) ACC and (B) DLPFC left. The colors indicate the areas covered by the subjects individually placed MRS voxels. The individual voxels were standardized with SPM12, overlapped in MRIcroGL and visualized in FSLeyes. ACC, anterior cingulate cortex; DLPFC L, left dorsolateral prefrontal cortex

### 2.4 ^1^H-MRS Analysis

The MRS data were analyzed using Osprey version 2.4.0, an all-in-one software for state-of-the-art processing and quantitative analysis of in vivo magnetic resonance spectroscopy (MRS) data(60). To optimally fit our varying TE, we simulated different basis sets for each TE (37–41), with a bandwidth of 2000Hz and 1024pts using MARSS in INSPECTOR version 11-2021(61). Our basis set included the following metabolites, based on recommendations in the LCModel manual(62): alanine, aspartate, creatine, GABA, glucose, glutamate, glutamine, glutathione, glycerophosphocholine, lactate, myoinositol, N-acetylaspartate, N-acetyl-aspartylglutamate, phosphocholine, phosphocreatine, scyllo-inositol and taurine. We added the default macromolecular and lipid components provided by Osprey to this basis set. For further analyses, we used the tissue and relaxation corrected molal concentration according to the Gasparovic method(63).

All spectra were visually inspected for artifacts. An exemplary fit of the spectra can be seen in Figure 1. Spectral exclusion criteria were, therefore, either visual failure of the fitting algorithm, a resultant FWHM>13 Hz in the ACC(64), or a CRLB>20% of glutamate+glutamine (Figure 1) concentration. The fits of the spectra were appropriate for all participants except for one, where the Glx concentration of the left DLPFC could not be estimated.

### 2.5 Resting-state fMRI Analysis

Functional images were pre-processed and denoised using the default options in CONN version 21a(65). Based on the subject-level quality control measures, we excluded eight participants. Details for the preprocessing, denoising and quality control can be found in the supplementary material.

We used the CONN-predefined language network voxels left pSTG and left IFG, and the left pMTG as our regions of interest to conduct the seed-based connectivity maps. The second-level analyses were then performed for the 46 subjects corrected for sex and age for our selected seeds (left IFG, left pSTG, left MTG). The significance level was set to a combination of a cluster-forming p<0.001 voxel-level threshold, and a familywise corrected p-FDR<0.05 cluster-size threshold. For further statistical analyses, we extracted beta values, representing the connectivity value between the cluster and the seed region, for each participant and seed region.

### 2.6 Computational analysis of the predictive language task

To analyze parameters from the predictive language task, we used a mechanistic Bayesian belief updating model to estimate the prior weight, which indicates the participants’ reliance on prior knowledge versus sensory input. The model dynamically updates the probability of a correct response based on prior context and degraded sensory evidence, following a beta-Bernoulli framework (see details in the supplementary materials or recent study(18)). For further analyses, we used the schizotypy-informed prior weight, which was simulated using normalized and centralized sample-averaged parameters for entropy, channel number, cloze probability, and clarity rating. With this approach, we could qualify the weighting of prior beliefs relative to the sensory likelihood and establish a personalized prior weight based on individual schizotypal levels. Furthermore, we calculated the proportion of TBH during the task, which refers to instances where participants perceived a word differently from what was presented.

### 2.7 Ridge regression for predictor identification of the schizotypy-informed prior weight, TBH, and schizotypy

We intended to examine associations between seed-based clusters and the schizotypy-informed prior weight, TBH, or schizotypy using linear regression, but the combination of 45 participants and up to 8–9 predictors per model posed a substantial risk of overfitting. We, therefore, first applied ridge regression (glmnet package version 4.1-8)(66), implemented using R Statistical Software (version 4.2.2)(67) to select the three most important predictors for each linear model. These “ridge”- models included the schizotypy-informed prior weight, the TBH, or the schizotypy (in analysis referred to as SPQ score) as outcome variables with the significant clusters (extracted betas values) from the second-level fMRI-analyses for each seed region (left IFG, left pSTG, left pMTG) entered as predictors. We limited the models to three clusters as predictors because we were specifically interested in their interaction with the two Glx measures which then resulted in five predictors in total, which suitable for 45 participants. See visualizations of the ridge regression in the supplementary material (Figure S5). The significance level was set to p<0.05. Analyses were visualized in R using the ggplot2 package version 3.4.1(68).

### 2.8 Interaction between FC and Glx as predictors of the schizotypy-informed prior weight, TBH, and schizotypy

Using the reduced set of predictors, we then ran linear models, which included the interaction between each FC cluster (extracted betas values) and Glx levels in the ACC and left DLPFC, resulting in five predictors each. The outcome variables were the schizotypy-informed prior weight, the TBH frequency, or schizotypy (SPQ score). All variables were standardized, and significance was set at p<0.05. A visual overview of our analysis can be found in the supplements (Figure S1).

## 3 Results

### 3.1 Significant connections of the language network

Figure 2 shows all significant clusters from the three seed regions within the language network identified in the second-level fMRI-analysis. Eight clusters significantly and positively correlated with the left IFG. Seven clusters were positively associated with the pSTG and one cluster correlated negatively. Seven clusters were positively associated with the left pMTG, while two clusters correlated negatively. Tables S2-S4 contain a detailed description of each significant cluster and its detailed location.

**Figure 2:**
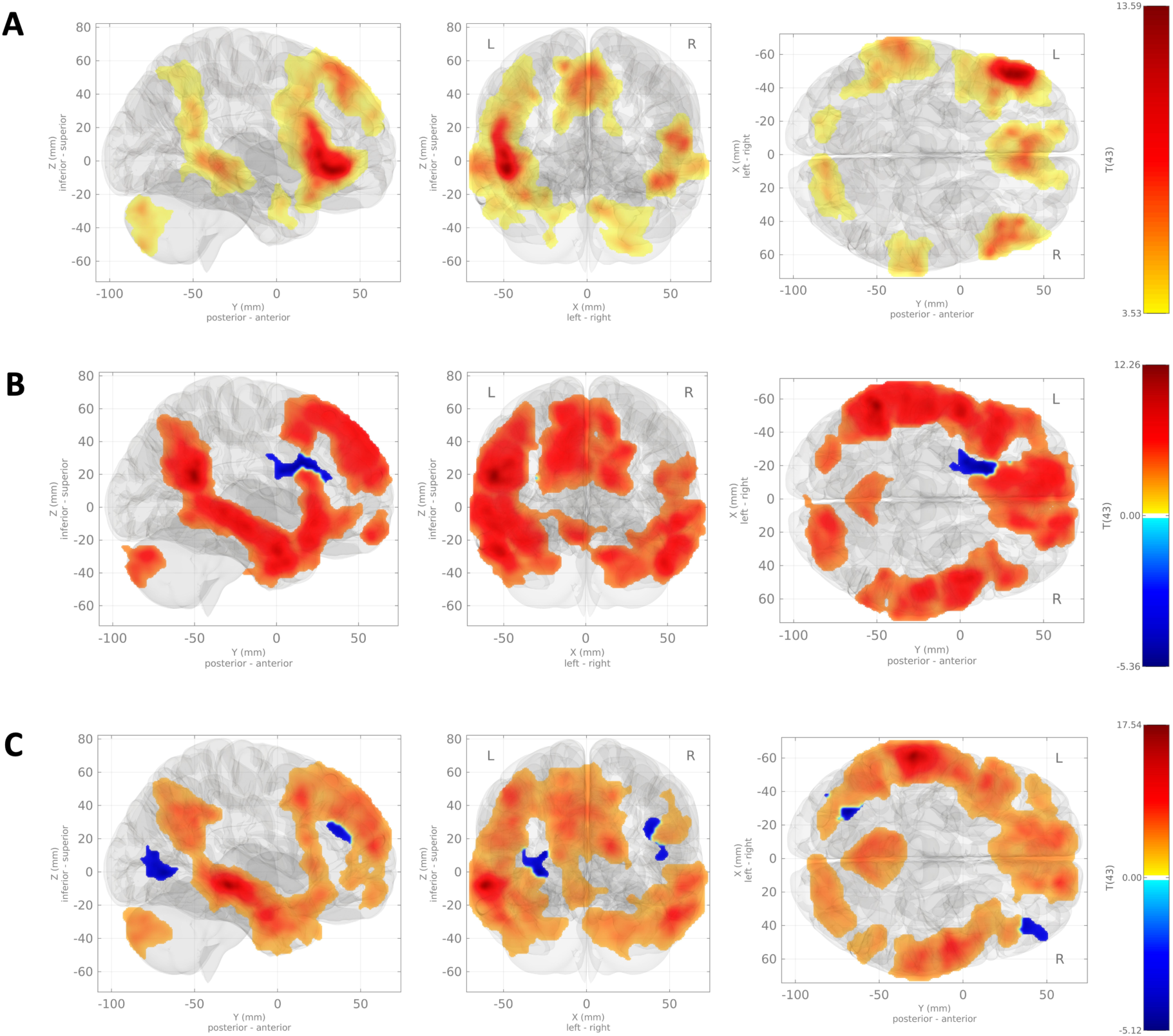
Overview of the results of second-level analysis. Note: 3D plots of significant clusters in our second level analysis: Effect of all subjects corrected for sex and age for A) left IFG, B) left pSTG, and C) left MTG. The significance level was set to voxel p uncorrected < 0.05 and Cluster p-FWE corrected < 0.001. IFG, inferior frontal gyrus; pSTG, posterior superior temporal gyrus; pMTG, posterior middle temporal gyrus

**Figure 3:**
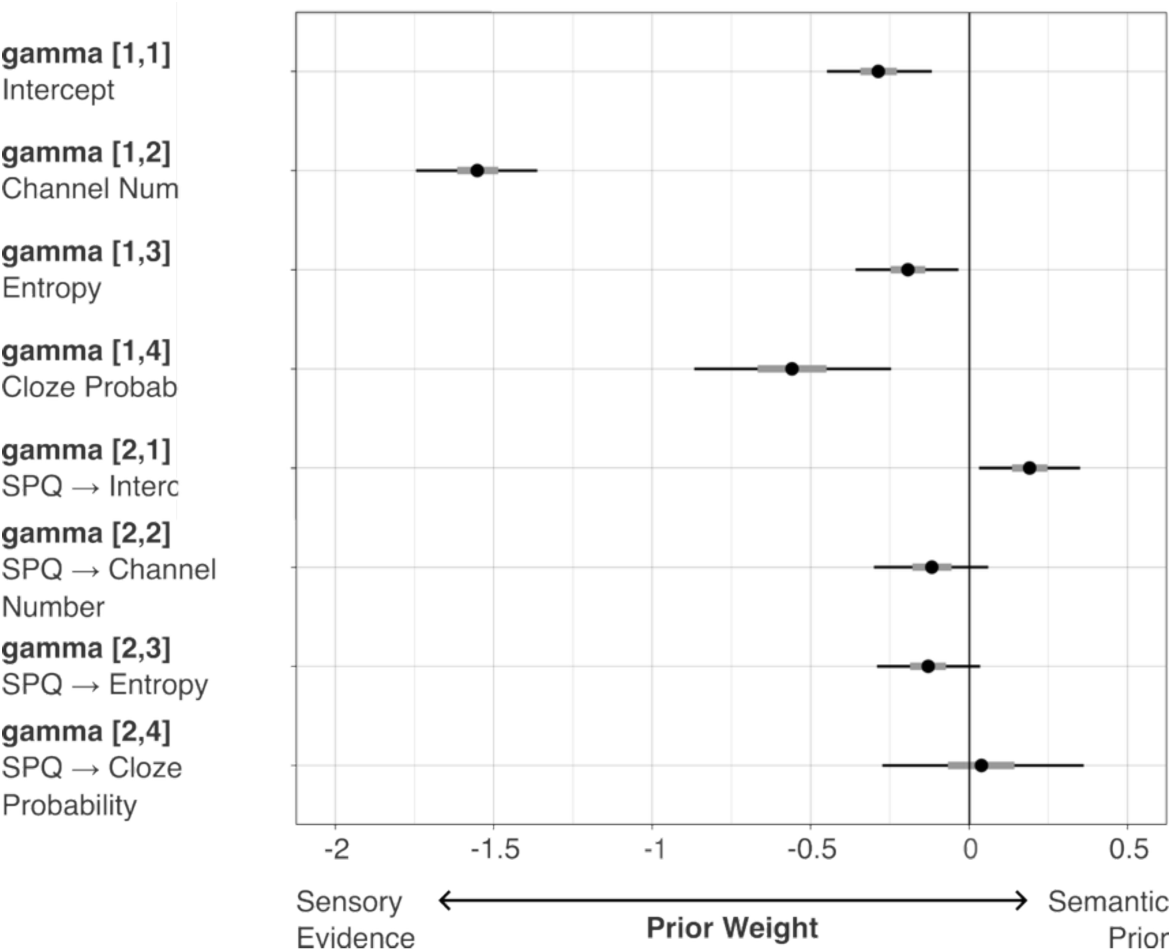
Impact of schizotypy on the prior weight. Note: For each trial of our task in Study 1 (N=53, left), we estimated the prior weight 𝜈, which represents how much the prior is weighted relative to the sensory evidence using the normative intra-trial Bayesian belief update. Simultaneously, to explain trial-level values of 𝜈, we used a regularized hierarchical linear model with task conditions as predictors and coefficients modulated by the participant’s SPQ score. Parameters with 95% CI excluding 0 indicate substantial contribution. For example, coefficient [gamma[1,2] describes how changes in acoustic degradation affect the prior weight. When the auditory input becomes clearer (i.e., as channel number increases and acoustic degradation decreases) individuals rely more on the sensory input, which is the expected pattern. Importantly, the SPQ-dependent modulation of the intercept (coefficient [gamma[2,1]]) quantifies how increasing schizotypy shifts the relative weight of the prior.

### 3.2 Levels of glutamate

The tissue and relaxation corrected molal concentration levels of Glx and their according quality parameters can be found in Table 1. Furthermore, in a previous paper(52) we described the association between Glx levels and schizotypy. We found that Glx concentrations in the ACC, but not in the left DLPFC, predicted schizotypy in healthy individuals.

**Table 1:**
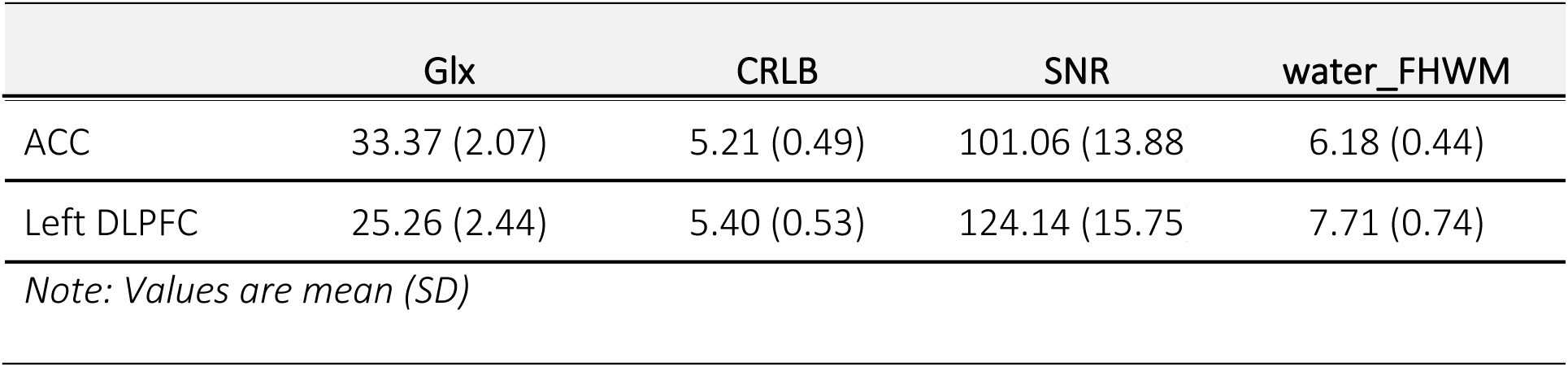
Glutamate levels and quality parameters.

| Glx | CRLB | SNR | water_FHWM |
| --- | --- | --- | --- |

|  |  |  |  |  |
| --- | --- | --- | --- | --- |
| ACC | 33.37 (2.07) | 5.21 (0.49) | 101.06 (13.88) | 6.18 (0.44) |
| Left DLPFC | 25.26 (2.44) | 5.40 (0.53) | 124.14 (15.75) | 7.71 (0.74) |

*Note: Values are mean (SD)*

### 3.3 Language task results

Our belief updating model estimation successfully converged and passed all quality control checks (see supplementary Figure S6). We observed a significant effect of schizotypy on the prior weight 𝜈 (Table 2: Parameter estimates for gammas for each task parameter indicating the population effect of SPQ (N=53), suggesting that individuals with higher levels of schizotypy assign greater weight toward prior beliefs over sensory evidence (coefficient gamma[2,1]). More detailed, our modelling results indicated that an increase of schizotypy (SPQ) by 10 points on the 5-point Likert-scale version of the SPQ (range: 0 - 296) is associated with a 3.8% increase in prior weight relative to sensory evidence (i.e., exp(−0.29+0.19/49.85)/exp(−0.29)=1.0038, where 49.85 is SPQ SD, -0.29 is the gamma[1,1] intercept, and 0.19 is gamma[2,1], reflecting SPQ impact). Posterior predictive simulations demonstrated the influence of increasing schizotypy on prior weight in typical trials (see supplementary Figure S7).

**Table 2:** Parameter estimates for gammas for each task parameter indicating the population effect of SPQ (N=53)

|  | Median | MAD | CI<br>2.5% | CI<br>25 % | CI<br>50 % | CI<br>75 % | CI<br>97.5% | n_eff | Rhat |
| --- | --- | --- | --- | --- | --- | --- | --- | --- | --- |
| gamma[1,1] (intercept) | -0.29 | 0.09 | -0.45 | -0.34 | -0.29 | -0.23 | -0.12 | 5415 | 1 |
| gamma[1,2]<br>(channel number) | -1.55 | 0.10 | -1.75 | -1.62 | -1.55 | -1.49 | -1.36 | 7650 | 1 |
| gamma[1,3] (entropy) | -0.19 | 0.08 | -0.36 | -0.25 | -0.19 | -0.14 | -0.03 | 8195 | 1 |
| gamma[1,4]<br>(cloze probability) | -0.56 | 0.16 | -0.87 | -0.67 | -0.56 | -0.45 | -0.25 | 7138 | 1 |
| gamma[2,1]<br>(SPQ → Intercept) | 0.19 | 0.08 | 0.03 | 0.14 | 0.19 | 0.25 | 0.35 | 5885 | 1 |
| gamma[2,2] (<br>SPQ → channel number) | -0.12 | 0.09 | -0.30 | -0.18 | -0.12 | -0.06 | 0.06 | 7741 | 1 |
| gamma[2,3]<br>(SPQ → entropy) | -0.13 | 0.08 | -0.29 | -0.19 | -0.13 | -0.07 | 0.04 | 9024 | 1 |
| gamma[2,4]<br>(SPQ → cloze probability) | 0.04 | 0.16 | -0.27 | -0.07 | 0.04 | 0.14 | 0.36 | 6803 | 1 |

*Note: In this model the prior weight was stratified by the effect of the clarity rating. As the posterior gamma distribution of clarity is uninterpretable, it is not presented in the table. MAD: Median absolute deviation; n\_eff: effective sample size, of 10000 samples have been drawn; Rhat: potential scale reduction factor on split chains, Rhat<1.04 indicates convergence*

### 3.4 Significant associations between ACC Glx and the schizotypy-informed prior weight

Detailed information on the linear model results can be found in Table 3–Table 5. In the final linear models, ACC Glx emerged as a significant predictor for the schizotypy-informed prior weight either as a main effect or in interaction with FC clusters. In the left pSTG model, ACC Glx significantly predicted the schizotypy-informed prior weight (β=0.39 ± 0.16, t=2.42, p=0.021). In contrast, for the left IFG seed, only the interaction between ACC Glx and the connectivity between the left IFG and IFG_L_Cluster_1 was significant (β=-0.79 ± 0.22, t=-3.53, p=0.001). IFG_L_Cluster_1 covered 93% of the pars opercularis and 81% of the pars triangularis, as well as 45% of the frontal orbital cortex and 35% of the left middle frontal gyrus.

**Table 3:**
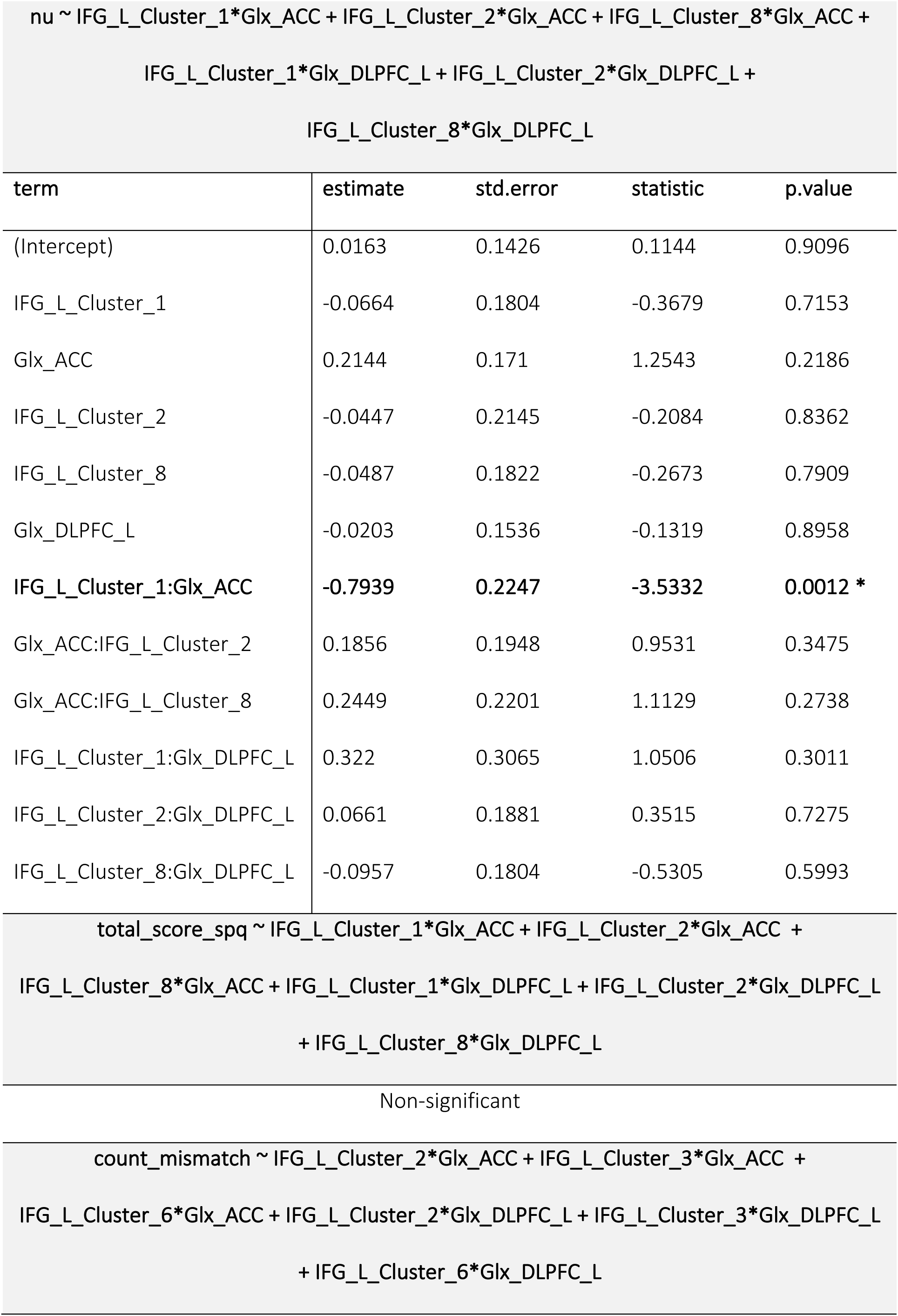
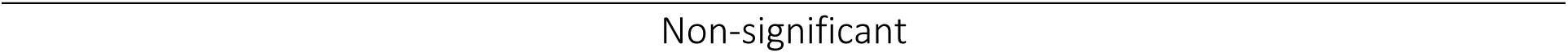
Significant linear models with left IFG as seed.

**Table 4:** Significant linear models with left pSTG as seed.

| nu ~ pSTG_L_Cluster_2*Glx_ACC + pSTG_L_Cluster_7*Glx_ACC +<br>pSTG_L_Cluster_8*Glx_ACC + pSTG_L_Cluster_2*Glx_DLPFC_L +<br>pSTG_L_Cluster_7*Glx_DLPFC_L + pSTG_L_Cluster_8*Glx_DLPFC_L |  |  |  |  |
| --- | --- | --- | --- | --- |
| term | estimate | std.error | statistic | p.value |
| (Intercept) | -0.0389 | 0.164 | -0.2374 | 0.8139 |
| pSTG_L_Cluster_2 | -0.1787 | 0.275 | -0.6497 | 0.5204 |
| Glx_ACC | **0.3854** | **0.1593** | **2.4194** | **0.0212 \*** |
| pSTG_L_Cluster_7 | **-0.6031** | **0.2206** | **-2.7343** | **0.01 \*** |
| pSTG_L_Cluster_8 | 0.0945 | 0.2252 | 0.4195 | 0.6775 |
| Glx_DLPFC_L | -0.1969 | 0.1672 | -1.1776 | 0.2474 |
| pSTG_L_Cluster_2:Glx_ACC | 0.0774 | 0.1981 | 0.3906 | 0.6986 |
| Glx_ACC:pSTG_L_Cluster_7 | 0.0587 | 0.2578 | 0.2279 | 0.8212 |
| Glx_ACC:pSTG_L_Cluster_8 | 0.3152 | 0.2083 | 1.5135 | 0.1397 |
| pSTG_L_Cluster_2:Glx_DLPFC_L | -0.3139 | 0.3415 | -0.9191 | 0.3647 |
| pSTG_L_Cluster_7:Glx_DLPFC_L | -0.2738 | 0.2148 | -1.2743 | 0.2115 |
| pSTG_L_Cluster_8:Glx_DLPFC_L | -0.1614 | 0.2612 | -0.6179 | 0.5409 |
| total_score_spq ~ pSTG_L_Cluster_2*Glx_ACC + pSTG_L_Cluster_3*Glx_ACC +<br>pSTG_L_Cluster_7*Glx_ACC + pSTG_L_Cluster_2*Glx_DLPFC_L +<br>pSTG_L_Cluster_3*Glx_DLPFC_L + pSTG_L_Cluster_7*Glx_DLPFC_L |  |  |  |  |
| term | estimate | std.error | statistic | p.value |

| (Intercept) | -0.2603 | 0.1651 | -1.5765 | 0.1244 |
| --- | --- | --- | --- | --- |
| Glx_ACC | 0.2858 | 0.1626 | 1.7571 | 0.0882 |
| pSTG_L_Cluster_2 | 0.0058 | 0.2015 | 0.0286 | 0.9773 |
| pSTG_L_Cluster_3 | 0.1728 | 0.1735 | 0.9962 | 0.3264 |
| pSTG_L_Cluster_7 | -0.3307 | 0.2058 | -1.6067 | 0.1176 |
| Glx_DLPFC_L | -0.0514 | 0.161 | -0.3191 | 0.7517 |
| Glx_ACC:pSTG_L_Cluster_2 | **0.4858** | **0.2077** | **2.3388** | **0.0256 \*** |
| Glx_ACC:pSTG_L_Cluster_3 | -0.4524 | 0.1794 | -2.5218 | **0.0167 \*** |
| Glx_ACC:pSTG_L_Cluster_7 | 0.5207 | 0.2507 | 2.0773 | **0.0456 \*** |
| pSTG_L_Cluster_2:Glx_DLPFC_L | -0.6338 | 0.2496 | -2.5394 | **0.016 \*** |
| pSTG_L_Cluster_3:Glx_DLPFC_L | 0.2761 | 0.1801 | 1.5334 | 0.1347 |
| pSTG_L_Cluster_7:Glx_DLPFC_L | -0.5007 | 0.2101 | -2.3832 | **0.0231 \*** |
| count_mismatch ~ pSTG_L_Cluster_1*Glx_ACC + pSTG_L_Cluster_3*Glx_ACC +<br>pSTG_L_Cluster_6*Glx_ACC + pSTG_L_Cluster_1*Glx_DLPFC_L +<br>pSTG_L_Cluster_3*Glx_DLPFC_L + pSTG_L_Cluster_6*Glx_DLPFC_L |  |  |  |  |
| term | estimate | std.error | statistic | p.value |
| (Intercept) | -0.0392 | 0.1606 | -0.2442 | 0.8086 |
| pSTG_L_Cluster_1 | -0.2862 | 0.2225 | -1.2864 | 0.2072 |
| Glx_ACC | -0.1528 | 0.177 | -0.8633 | 0.3942 |
| pSTG_L_Cluster_3 | 0.1576 | 0.2027 | 0.7776 | 0.4423 |
| **pSTG_L_Cluster_6** | **0.463** | **0.1983** | **2.3347** | **0.0258 \*** |
| Glx_DLPFC_L | 0.0848 | 0.1637 | 0.518 | 0.6079 |
| pSTG_L_Cluster_1:Glx_ACC | 0.0489 | 0.2393 | 0.2045 | 0.8392 |
| Glx_ACC:pSTG_L_Cluster_3 | -0.1165 | 0.2369 | -0.4915 | 0.6263 |
| Glx_ACC:pSTG_L_Cluster_6 | 0.1592 | 0.1634 | 0.9747 | 0.3368 |
| pSTG_L_Cluster_1:Glx_DLPFC_L | -0.385 | 0.3001 | -1.2829 | 0.2084 |
| pSTG_L_Cluster_3:Glx_DLPFC_L | 0.2566 | 0.2009 | 1.277 | 0.2105 |
| pSTG_L_Cluster_6:Glx_DLPFC_L | 0.0272 | 0.2459 | 0.1105 | 0.9127 |

**Table 5:**
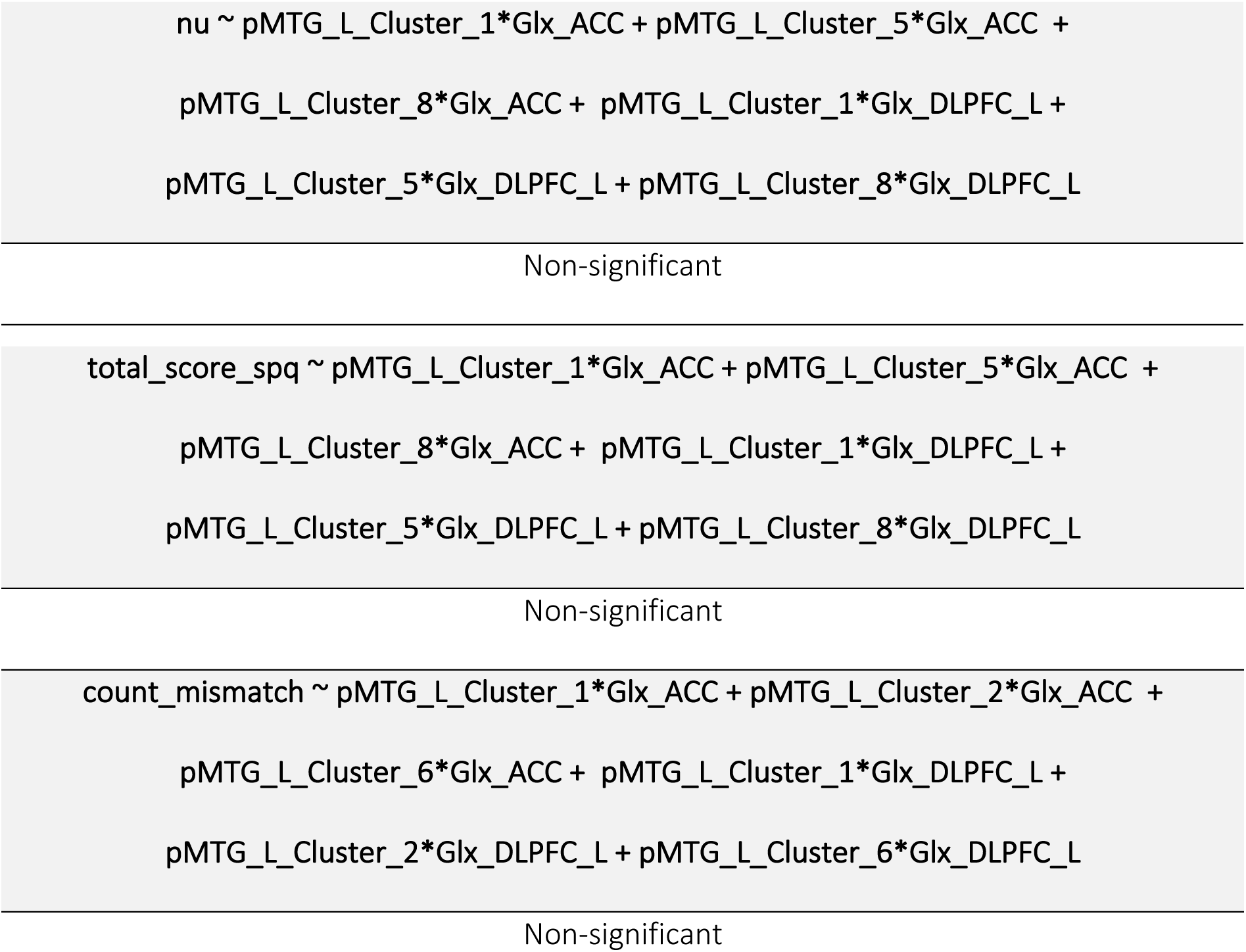
Significant linear models with left pMTG as seed.

These findings imply that higher ACC Glx concentrations are associated with greater prior overweighting during predictive sentence processing in individuals with increasing schizotypy. In contrast, under lower ACC Glx concentrations, stronger connectivity between the left IFG and frontal areas is linked to greater prior overweighting during predictive sentence processing in individuals with increasing schizotypy.

### 3.5 FC predicting the schizotypy-informed prior weight

Moreover, we found that the connectivity between the left pSTG and pSTG_Cluster_7 significantly and negatively predicted the schizotypy-informed prior weight (nu; β=-0.60 ±0.22, t=-2.73.19, p=0.010). This cluster included parts of the left corpus callosum. This finding suggests that more negative connectivity between the left pSTG and corpus callosum is associated with greater prior overweighting during predictive sentence processing in individuals with increasing schizotypy.

### 3.6 Significant associations between FC of the left pSTG and TBH

TBH were significantly predicted by the connectivity between the pSTG and pSTG_L_Cluster_6 (β=0.46 ± 0.20, t =2.34, p=0.026), which covered the left frontal pole (5%) and frontal medial cortex (1%). This association indicated that with increasing connectivity between the left pSTG and left frontal pole, individuals experienced more TBH.

### 3.7 FC predicting schizotypy by interaction with Glx levels

Schizotypy was significantly predicted by the interaction between ACC Glx and pSTG_L_Cluster_2 (β=0.49 ± 0.21, t=2.34, p=0.026), pSTG_L_Cluster_3 (β=-0.45 ±0.18, t=-2.52, p=0.017), as well as the pSTG_L_Cluster_7 (β=0.52 ±0.25, t=2.08, p=0.046). pSTG_L_Cluster_2 included 40% of the left and 22 % of the right superior frontal gyrus (SFG) as well as 21% of the left frontal pole, pSTG_L_Cluster_3 mainly covered the right MTG (91% of anterior and 67% of posterior division) and right anterior inferior temporal gyrus (aITG, 63%), whereas pSTG_L_Cluster_7 included parts of the left Corpus Callosum. The results suggest that higher ACC Glx enhances the influence of pSTG connectivity on schizotypy, increasing schizotypy when the connectivity of the left pSTG with clusters 2 and 7 is stronger, but decreasing it when connectivity with cluster 3 is stronger.

Furthermore, we found a significant, negative interaction effect between left DLPFC Glx and pSTG_L_Cluster_2 (frontal areas, β=-0.63 ±0.25, t=-2.54, p=0.016) as well as pSTG_L_Cluster_7 (parts of the left Corpus Callosum, β=-0.50 ±0.21, t=-2.38, p=0.023). This implies that under higher left DLPFC Glx concentrations, stronger connectivity between the left pSTG and these clusters is associated with lower schizotypy, suggesting an inhibitory or modulatory role of DLPFC glutamate on the relationship between pSTG connectivity and schizotypal traits.

We did not find linear associations between the prior weights of the different conditions and the significant clusters for the significant clusters of the left pMTG.

## Discussion

The goal of this study was to explore whether, how, and to what extent the interplay between resting-state FC in the language network and cortical levels of Glx predict imbalances in prior weighting during language processing, as well as the occurrence of TBH and schizotypal traits. Our findings revealed three key results: (1) the overweighting of schizotypy-informed semantic priors during language processing was significantly predicted by ACC Glx and by an interaction ACC Glx with an increased FC between the left IFG, and the inferior and middle frontal areas; (2) the frequency of TBH during the task was associated with increased connectivity between the left pSTG and medial frontal areas; and (3) schizotypy was predicted by interactions between ACC and left DLPFC Glx levels with FC between the left pSTG and superior, middle, and inferior frontal brain areas. These findings provide novel insights into the neurobiological and computational mechanisms underlying subclinical psychotic symptoms.

The impact of ACC Glx and resting-state FC on the schizotypy-informed prior weighting This study extends our previous findings(18) on the role of ACC Glx in the overweighting of priors by linking it to resting-state FC of language regions. The schizotypy-informed prior weighting was predicted by ACC Glx as a main effect in the left pSTG analysis and in the left IFG model by the interaction between ACC Glx and functional connectivity to frontal areas. This suggests a general modulatory role of ACC glutamate on frontal connectivity which may contribute to alterations in the prior weighting. However, a plausible explanation for these different effects lies in the distinct computational roles of the left IFG and pSTG within the language network. The IFG is involved in integrating semantic information(69–73), and generating predictive inference in broader cognitive contexts(74–76), and therefore in top-down Processing within the cortical hierarchy(4). In contrast, the pSTG seems to be more strongly associated with sensory-driven, bottom-up analyses and prediction-error evaluation during speech and auditory processing(77–79). Therefore, the absence of an ACC Glx x pSTG-FC interaction on the schizotypy-informed prior weighting may indicate that ACC-glutamatergic modulation preferentially influences regions involved in generative, top-down processing, such as the IFG. Furthermore, previous studies have shown that ACC glutamate excitation appears to relate to a stronger reliance on prior beliefs(18,80). This is consistent with the main effect observed in the pSTG connectivity model, where ACC Glx alone predicted higher prior weighting.

Interestingly, we observed a negative interaction between the ACC Glx and an IFG-frontal FC, such that higher ACC Glx in combination with a lower IFG–frontal FC was associated with stronger prior overweighting. This finding proposes that E/I-imbalances may shape large-scale network connectivity and thereby enhance reliance on priors. This interpretation also aligns with the heterogeneity reported in the existing literature, where studies showed mixed associations between ACC Glx and perceptual or cognitive processes(36,81,82). Cai et al.(82), for example, reported a positive correlation between ACC Glx and impaired sensory integration, whereas Levar et al.(81) found no association of ACC Glx itself, only in relation to GABA. In contrast, Overbeck et al.(36) reported negative associations between ACC Glx and connectivity with several regions, including the medial prefrontal cortex.

Overall, our results suggest that the observed FC patterns may reflect stronger top-down control during language processing, potentially influencing how prior beliefs are weighted in individuals with higher schizotypal traits(4). These effects were associated with ACC Glx, which may influence top-down computations via its role in regulating E/I-balance and NMDA-dependent predictive signaling in frontal cortical circuits(10,83–85). Impairments in these mechanisms and their effect on behavioral and computational outcomes in SSDs have been shown in previous studies(86–88).

In addition to these results, the left pSTG connectivity model showed that reduced connectivity between the left pSTG and the left Corpus Callosum predicted the prior weighting in individuals with increasing schizotypy. Given that callosal pathways support interhemispheric auditory and language integration(89), reduced connectivity in this region may indicate less efficient sensory integration. From a predictive-processing perspective, weakened interhemispheric exchange could reflect reduced sensory precision, which in turn increases reliance on priors. However, these mechanistic links require further investigation in future work.

### TBH enhance left pSTG- frontal connectivity

The frequency of TBH during the task was associated with enhanced resting-state FC between the left pSTG and a frontal cluster. This is consistent with the role of the STG in the formation of auditory verbal hallucinations (AVH). For example, Homan et al.(90) identified tonic hyperactivity in the left STG in patients with AVH, suggesting it may serve as a trait marker. Neurostimulation studies have further demonstrated that targeting the left pSTG reduces the severity and/or frequency of AVH in clinical populations(91–93). Notably, modulating cortical excitability with transcranial direct current stimulation in the left pSTG also influenced the frequency of auditory misperceptions of voices in healthy individuals(94), supporting the idea that similar mechanisms may underlie clinical and subclinical hallucinations. In addition to the pSTG, other temporal and frontal regions within the language network have been associated with AVH(48,95–97).

Our findings consistently show hyperconnectivity; this is in line with results reported in a meta-analysis(98) indicating that AVH are more commonly associated with increased rather than decreased activity in schizophrenia patients. Our results therefore suggest that the neurobiological mechanism associated with TBH may share certain features with those underlying clinical hallucinations. Accordingly, nonclinical hallucinations could serve as a valuable model for studying hallucinations or hallucination-related processes under controlled conditions, which could support efforts to identify predictive biomarkers and for evaluating the effectiveness of early interventions(99–101).

Increased schizotypal traits are associated with a left pSTG FC and Glx interaction The interaction between ACC and left DLPFC Glx levels with resting-state FC of the left pSTG and frontal regions predicted schizotypy, indicating that glutamatergic modulations of connectivity networks may already be detectable in subclinical stages. Studies in healthy individuals suggest that both glutamate and GABA influence connectivity patterns(102,103), offering a potential biochemical account for understanding the connectivity alterations observed in clinical populations. While for FC-glutamate associations in schizophrenia some studies report significant associations(36,104–107) and others failing to find a link(81,108–111), our findings showed that glutamatergic-FC interactions vary already with schizotypy, suggesting that these alterations may be already present before the emergence of clinical symptoms. This is consistent with the view that schizotypy represents a stable personality trait(112).

### No left MTG connectivity effects

Interestingly, we found no significant associations between the left MTG connectivity and the schizotypy-informed prior weight, TBH, or schizotypy. While the MTG has been described to contribute to hallucinations in patients(113,114) and to internal feedback control circuits for speech production(115), our findings are inconclusive. MTG-associate connectivity alterations may only develop in later stages of the disorder, which requires further investigations, especially longitudinal studies.

### Limitations and future directions

In the present study, we focused on three key hubs of the language network, selected because they are central to the mechanism engaged during our predictive language task. However, future studies should extend this approach to other networks, such as the default mode and salience networks, for a more comprehensive understanding. Additionally, the voxels used to assess glutamate concentration were not located in the same regions as the language seeds, limiting our ability to draw direct conclusions about glutamate levels in language-specific areas. Future research should explore glutamate and GABA levels in language regions using spectral editing techniques to better understand their interactions. Finally, while our findings highlight associations between glutamate, resting-state FC, and schizotypy, the methods used do not allow for causal inferences. Future studies employing longitudinal or interventional designs could help clarify the directionality of these relationships.

## Conclusion

This study investigated the interplay between resting-state FC in the language network and cortical glutamate levels in predicting weighting of prior beliefs during language processing, TBH, and schizotypy. Our findings suggest that ACC glutamatergic signaling may play a role in shaping how prior beliefs influence language processing in schizotypy, although the precise mechanisms will require further investigation. Furthermore, Glx in the ACC and left DLPFC seem to modulate the relationship between pSTG-frontal resting-state FC in predicting schizotypy, suggesting that these neurobiological alterations are present even in subclinical stages. TBH were associated with increased resting-state FC of the left pSTG to frontal areas. Employing a multimethod approach, this study provides insights into how alterations in the resting-state FC within the language network and glutamate levels may contribute to early indicators of SSDs, even in a healthy subclinical sample. Future studies should investigate the progression of these effects over time and their potential as predictive biomarkers.

## Data Availability

All data produced are available online at OSF (see link below).

https://osf.io/36wgj/overview?view_only=94c2bbfefc6a4d13b99abf0b9e17db8d

## Acknowledgments

We would like to thank all participants for their time and engagement.

## Author contributions: CRediT

*Verena Demler:* Investigation, Data collection, Formal analysis, Visualization, Writing – original draft. *Elisabeth Sterner:* Investigation, Data collection, Writing – review and editing. *Franziska Knolle:* Conceptualization, Methodology, Validation, Writing – review and editing, Project administration, Supervision.

## Conflict of interest

The authors declare no conflict of interest.

Ethics: The medical research ethics committee of the Technical University of Munich gave ethical approval for this work. All subjects gave written informed consent in accordance with the Declaration of Helsinki.

## Data availability

Data is available upon reasonable request to the corresponding author.

