## Supplemental material for "Language-network connectivity and cortical glutamate and their associations with overweighted semantic prior beliefs, schizotypy and task-based hallucinations"

### 1 **Methods**

#### 1.1 **Participants**

**Inclusion criteria:** Native German speaker; right-handed; no diagnosis of schizophrenia, psychosis or autism, or any neurological disease or injury; not currently taking any psychoactive medication within at least the past six weeks and no contraindications for MRI scanning.

**Subjects:** Out of the participants, three had been previously diagnosed with depression, two had prior eating disorders, one was diagnosed with schizoid personality disorder along with adjustment disorder, and another reported having post-traumatic stress disorder and narcissistic personality disorder. However, only one of the participants was consistently taking antipsychotic medication (fluoxetine, atomoxetine) for attention deficit disorder.

Details of demographic data and symptom scores are shown in Table S1 and have been previously published (1–3).

#### 1.2 **MRS Data Acquisition and Analysis**

To collect the MRS data, we used a ¹H-MRS single voxel ECHO volume Point Resolved Spectroscopy Sequence (PRESS) sequence with the following scan parameters: TE set to shortest (resulting in a range between 37.3 ms and 41.2 ms, which is accounted for in the basis sets); TR, 2000ms; 16 phase cycle steps; acquisition BW, 2 kHz; 1024 data points; flip angle, 90°. Furthermore, we applied the conventional Philips water suppression technique (excitation), which performs Automatic Water Suppression Optimization (AWSO) pre-scans, to minimize residual water.

To fit and quantify our data, we used the LCModel (LCM) implementation in Osprey. The LCModel algorithm fits spectra in the frequency domain using a linear combination model (4). Our processing followed established parameters, featuring a metabolite fit range from 0.5 to 4.0 ppm and a water fit range from 2.0 to 7.4 ppm. A knot spacing of 0.4 ppm was applied. Furthermore, Osprey utilizes the SPM12 segmentation function to divide the structural image into tissue probability maps (5). These maps are then overlaid with the co-registered voxel masks created by the Co-registration module, enabling the calculation of fractional tissue volumes for gray matter (GM), white matter (WM), and cerebrospinal fluid (CSF). To estimate the absolute levels of Glx (glutamate+glutamine), Osprey combined with LCM calculates the tissue and relaxation corrected molal concentration (mol/kg) following the Gasparovic method (6).

#### 1.3 **Resting-state fMRI Acquisition and Processing**

Resting-state fMRI was acquired during the same session with a multiband-SENSE sequence using the following parameters: TR=2.3s; TE=30ms; SENSE acceleration factor=2; multiband factor=2; 250 time-points; slice thickness=2mm; slice gap=0.2mm; voxel size 2x2x2 mm; slices, 62; flip angle 75°; field of view 192x192mm; matrix size 96x94; total scan time 584s.

Results included in this manuscript come from analyses performed using CONN (7) (RRID:SCR_009550) release toolbox version 22a (8) and SPM(9) (RRID:SCR_007037) release 12.6225. The following methods part derives from the methods description provided by CONN.

**Preprocessing:** Functional and anatomical data were preprocessed using a flexible preprocessing pipeline (10) including realignment with correction of susceptibility distortion interactions, slice timing correction, outlier detection, direct segmentation and MNI-space normalization, and smoothing. Functional data were realigned using SPM realign & unwarp procedure (11), where all scans were coregistered to a reference image (first scan of the first session) using a least squares approach and a 6 parameter (rigid body) transformation (12), and resampled using b-spline interpolation to correct for motion and magnetic susceptibility interactions. Temporal misalignment between different slices of the functional data (acquired with multiband factor 2, FH order) was corrected following SPM slice-timing correction (STC) procedure (13,14), using sinc temporal interpolation to resample each slice BOLD timeseries to a common mid-acquisition time. Potential outlier scans were identified using ART (15) as acquisitions with framewise displacement above 0.9 mm or global BOLD signal changes above 5 standard deviations (16,17), and a reference BOLD image was computed for each subject by averaging all scans excluding outliers. Functional and anatomical data were normalized into standard MNI space, segmented into grey matter, white matter, and CSF tissue classes, and resampled to 2 mm isotropic voxels following a direct normalization procedure (17,18) using SPM unified segmentation and normalization algorithm (19,20) with the default IXI-549 tissue probability map template. Last, functional data were smoothed using spatial convolution with a Gaussian kernel of 6 mm full width half maximum (FWHM).

**Denoising:** In addition, functional data were denoised using a standard denoising pipeline (21) including the regression of potential confounding effects characterized by white matter timeseries (5 CompCor noise components), CSF timeseries (5 CompCor noise components), motion parameters and their first order derivatives (12 factors) (22), outlier scans (below 21 factors) (16), session and task effects and their first order derivatives (2 factors), and linear trends (2 factors) within each functional run, followed by bandpass frequency filtering of the BOLD timeseries (23) between 0.008 Hz and 0.09 Hz. CompCor (24,25) noise components within white matter and CSF were estimated by computing the average BOLD signal as well as the largest principal components orthogonal to the BOLD average, motion parameters, and outlier scans within each subject's eroded segmentation masks. From the number of noise terms included in this denoising strategy, the effective degrees of freedom of the BOLD signal after denoising were estimated to range from 76.6 to 84.5 (average 83.8) across all subjects (17).

**First-level analysis SBC_01:** Seed-based connectivity maps (SBC) were estimated, characterizing the patterns of functional connectivity with 169 ROIs; relevant for further analysis three ROIS from the language network (8). Functional connectivity strength was represented by Fisher-transformed bivariate correlation coefficients from a weighted general linear model (weighted-GLM (26)), defined separately for each pair of seed and target areas, modeling the association between their BOLD signal timeseries. In the order to compensate for possible transient magnetization effects at the beginning of each run, individual scans were weighted by a step function convolved with an SPM canonical hemodynamic response function and rectified.

**Group-level analyses** were performed using a General Linear Model (GLM (27)). For each individual voxel a separate GLM was estimated, with first-level connectivity measures at this voxel as dependent variables (one independent sample per subject) and groups (high-prior, low-prior) as independent variables. Voxel-level hypotheses were evaluated using multivariate parametric statistics with random-effects across subjects and sample covariance estimation across multiple measurements. Inferences were performed at the level of individual clusters (groups of contiguous voxels). Cluster-level inferences were based on parametric statistics from Gaussian Random Field theory (28,29). Results were thresholded using a combination of a cluster-forming p < 0.001 voxel-level threshold and a familywise corrected p-FDR < 0.05 cluster-size threshold (30).

1.1 **Computational analysis of the predictive language task**

First, we designed the predictive language task to reflect a Bayesian inference process with the sentence beginning reflecting the prior, and the cloze probability and the sensory degradation (i.e., channel number) of the sentence-final word capturing the sensory likelihood. The clarity rating was a measure for the precision of the prior weight. For the analysis of our parameters, we used a mechanistic Bayesian belief updating, which allowed us to estimate the prior weight, indicating how much participants relied on prior knowledge relative to the sensory input. This model operates on the assumption that participants strive to maximize the probability of providing a correct answer. In each experimental trial, the probability of correctness is updated dynamically: it begins with an initial (prior) value derived from the prior-inducing phrase and is then adjusted to a posterior value upon hearing the degraded target word, which serves as sensory evidence. This updating process follows the principles of Bayesian inference within a beta-Bernoulli model. The observation, being binary (correct or incorrect), is modeled using a Bernoulli distribution, while the prior and posterior distributions, representing uncertain probabilities of correctness, are modeled as beta distributions. According to Bayes’ rule, the update to the expected likelihood of correctness is computed as a precision-weighted prediction error. The prediction itself is defined as the cloze probability of the provided answer, representing what participants anticipate before encountering the final stimulus.  The prediction error then is the correspondence of the stimulus to the answer (1 or 0) minus the prediction. This error is then weighted by a factor of $1/(1+\nu)$, where $\nu$ is an implied number of previous observations capturing the relative strength or weight of the prior compared to sensory evidence (31). This strength $\nu$ of the prior was estimated on a trial-by-trial basis using a linear model that incorporated task-specific parameters such as entropy, the cloze probability of the stimulus (distinct from the cloze probability of the answer, which serves as the prior), channel number, and clarity rating as predictors. Additionally, schizotypy was included as a factor informing $\nu$, and highlighting its role in modulating the relative weight of prior information. A more detailed description of the task and the modeling can be found in (3).

For the analyses in this paper, we used the schizotypy-informed prior weight, which was simulated using normalized and centralized sample-averaged parameters for entropy, channel number, cloze probability, and clarity rating. With this simulation, we could qualify the overweighting of prior belief and establish a personalized prior weight based on individual schizotypal levels. Furthermore, we calculated the proportion of misperceptions (i.e., non-clinical task-based hallucinations) during the task, which refers to instances where participants perceived a word differently from what was actually presented.

### 2 **Results and Discussion**

#### 2.1 **Quality assessment rs-fMRI**

Quality control (QC) of our participant data was based on descriptions and recommendations of the paper of Morfini et al.(32). First, we did visual control of functional and structural normalization and segmentation by using preprocessing output plots from CONN, overlaying each participant’s anatomical and functional images. For example, in Figure S2 an outline of the MNI TPM template was added to the overlayed normalized functional data, showing a good overlap of our participants images with the template. CONN also produces several automated QC summary measures such as Invalid Scans, Proportion of Valid scans (PVS), and Mean Motion. The total number of scans identified as outliers (InvalidScans) and the ratio between non-outlier scans to all scans (PVS) serve as important indicators of the overall quality and quantity of valid data obtained for every subject. On the other hand, the mean motion - the average of the FD time series, calculated only over non-outlier scans - is more representative of the state of the data after pre-processing. In the context of QC summary measures, extreme values were defined as those exceeding or reaching the threshold Q3 + 3 interquartile range (IQR) or falling below Q1–3 IQR (see Figure S3).

Finally, the QC-FC measure in % (percent match in QC-FC correlations) examines whether alterations in the spatial correlation structure of the BOLD data were associated with participant-level quality control measures and therefore serves to evaluate the quality control of the entire dataset, rather than the quality of individual participants or runs. The QC-FC % values were used to assess whether the preprocessing and denoising steps resulted in satisfactory fMRI data quality levels. Match levels above 95% indicated minimal changes in the BOLD signal correlation structure, while lower values indicated potential issues in the denoised data, requiring alternative preprocessing or stricter participant exclusion criteria. In Figure S4 you can see that for the individual plots (Figure S4, B-G) the QC-FC % values are as recommended above the cut-off value of 95%, just for the overall plot the threshold was not quite reached (Figure S4, A)

#### 2.2 **Second-level analysis**

We performed second-level analyses for 46 subjects corrected for sex and age for our selected seeds (left IFG, left pSTG, left MTG). The significance level was set to cluster-forming p < 0.001 voxel-level threshold and a familywise corrected p-FDR < 0.05. The significant clusters are not pre-described brain regions but comprise a proportion of these. The composition of each Cluster can be found in Table S2 for the left IFG, Table S3 for the left pSTG and Table S4 for the left pMTG.

#### 2.3 **Ridge Regression and Linear Models**

We used ridge regression to identify the three most important predictors of our linear models, where we wanted to have a look at the effect of language connectivity and glutamate on the weighting of prior and on schizotypy. In Figure S5, you can see which three predictors had the biggest influence on the model for each linear model and were therefore chosen for the smaller linear model. These three predictors were then used together with an interaction with both Glx levels in the ACC and left DLPFC for the final linear models. The results of the linear models are shown in Table 3 in the manuscript for the left IFG, in Table 4 for the left pSTG, and in Table 5 for the left pMTG as seed region of or resting state analysis.

### 3 **Figures**

Figure S1: Overview of study design and analysis

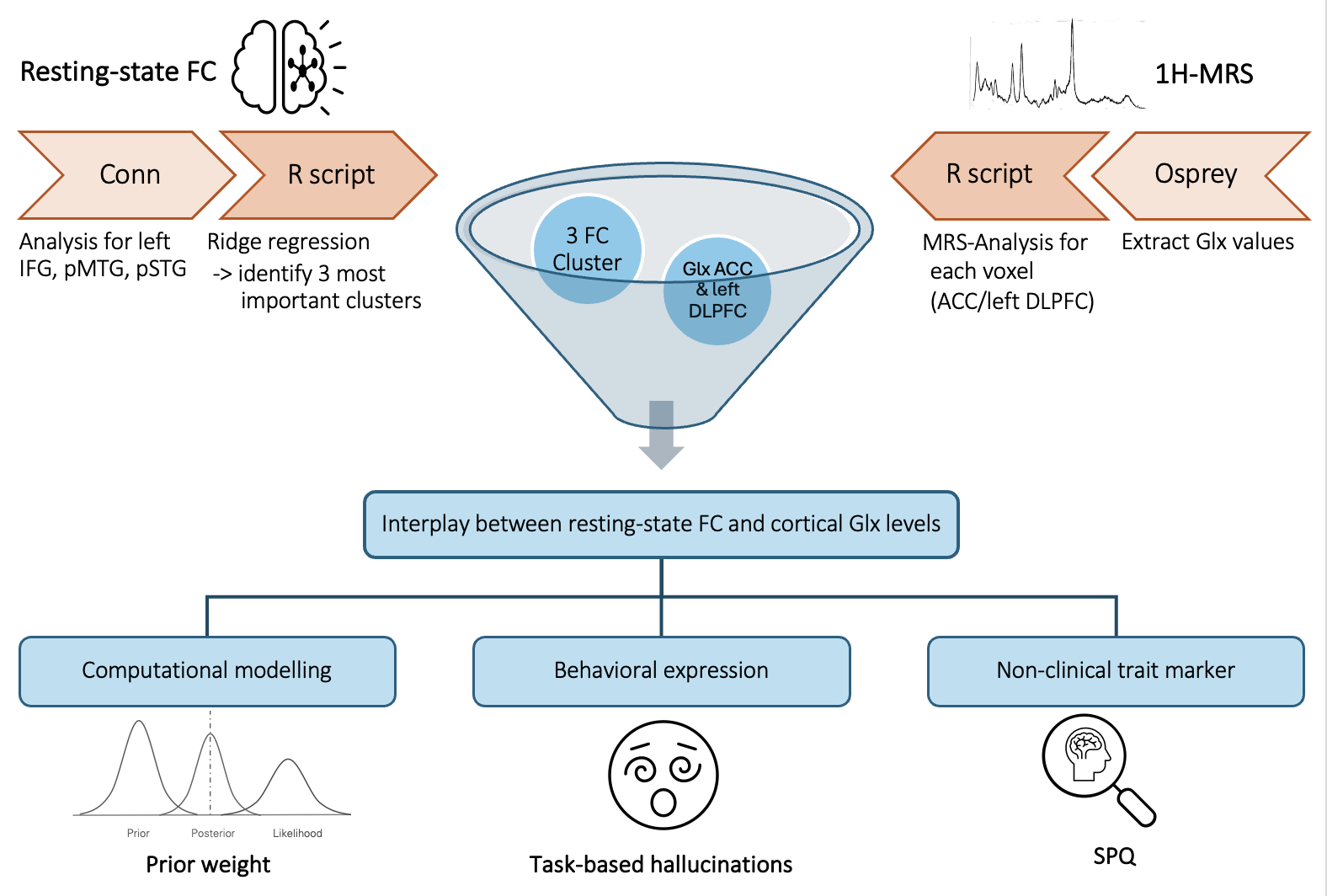

*Note: Visual overview of our study design. For detailed information on each step, refer to the methods section of our paper and supplements.*

Figure S2: Quality control of normalization of functional data + outline of MNI TPM template (46)

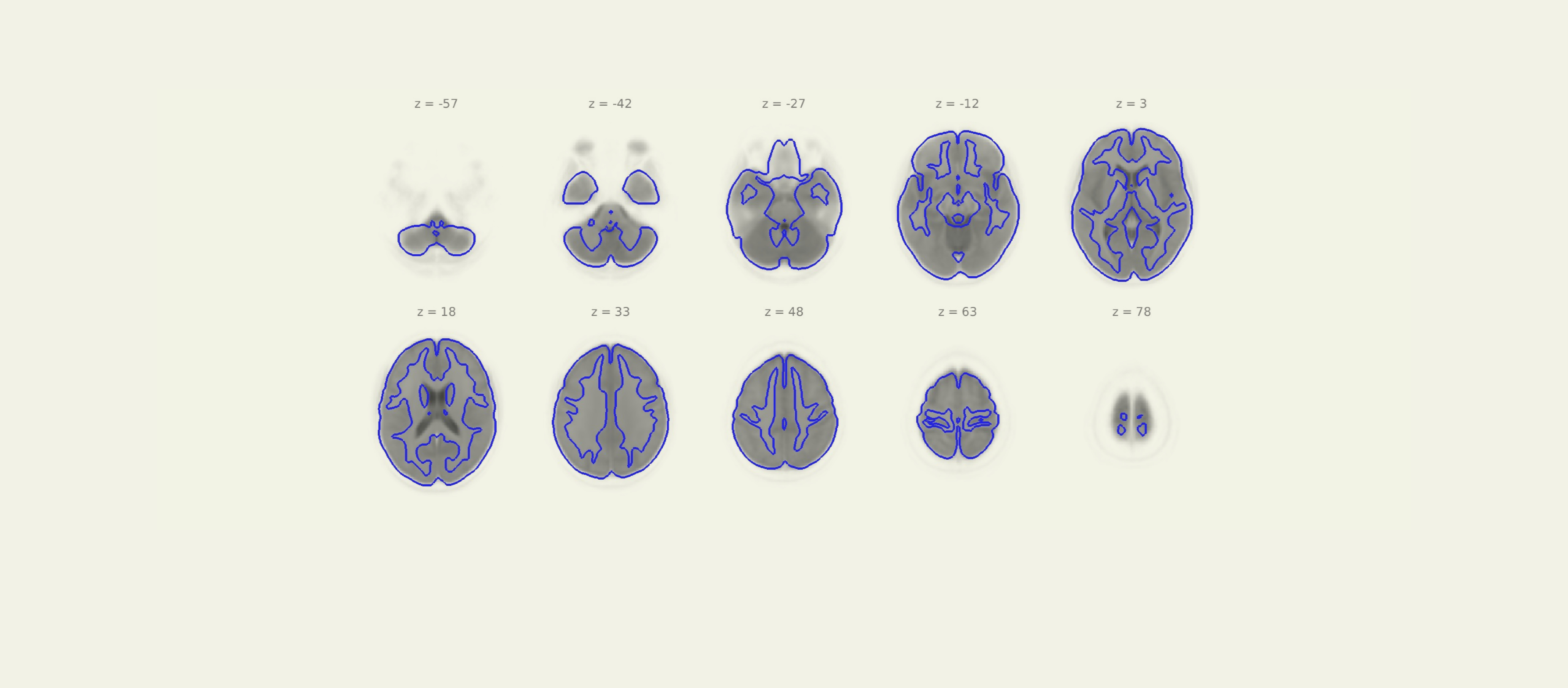

*Note: CONN’s preprocessing output plot shows an outline of the MNI TPM template, added on an overlay of each participant’s functional images.*

Figure S3: Distribution of subject-level Quality Control measures (53)

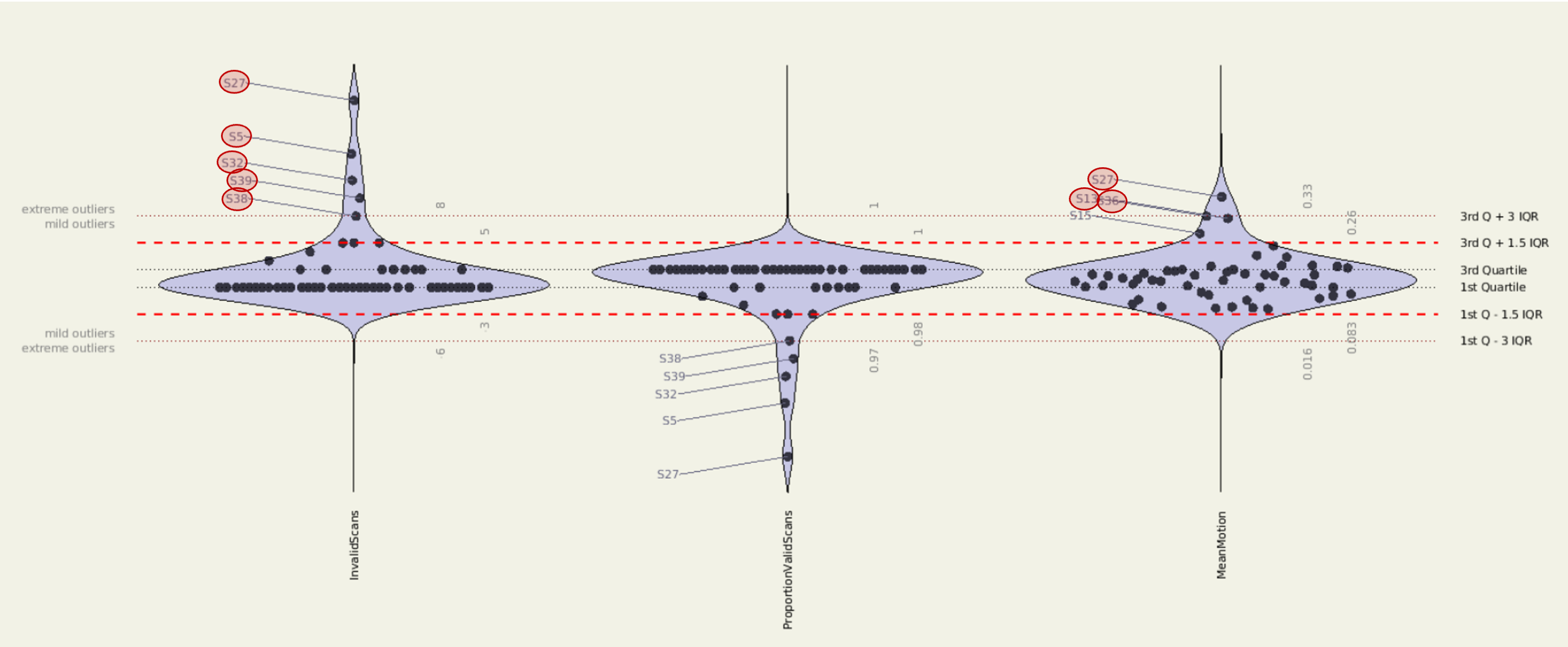

*Note: Distribution of tree subject level QC measures: Invalid Scans, the Proportion of Invalid Scans and Mean Motion (from left to right); extreme values, marked with red circles, were defined as those exceeding or reaching the threshold Q3 + 3 interquartile range (IQR) or falling below Q1–3 IQR*

Figure S4: Distribution of QC-FC associations (QC-FC)

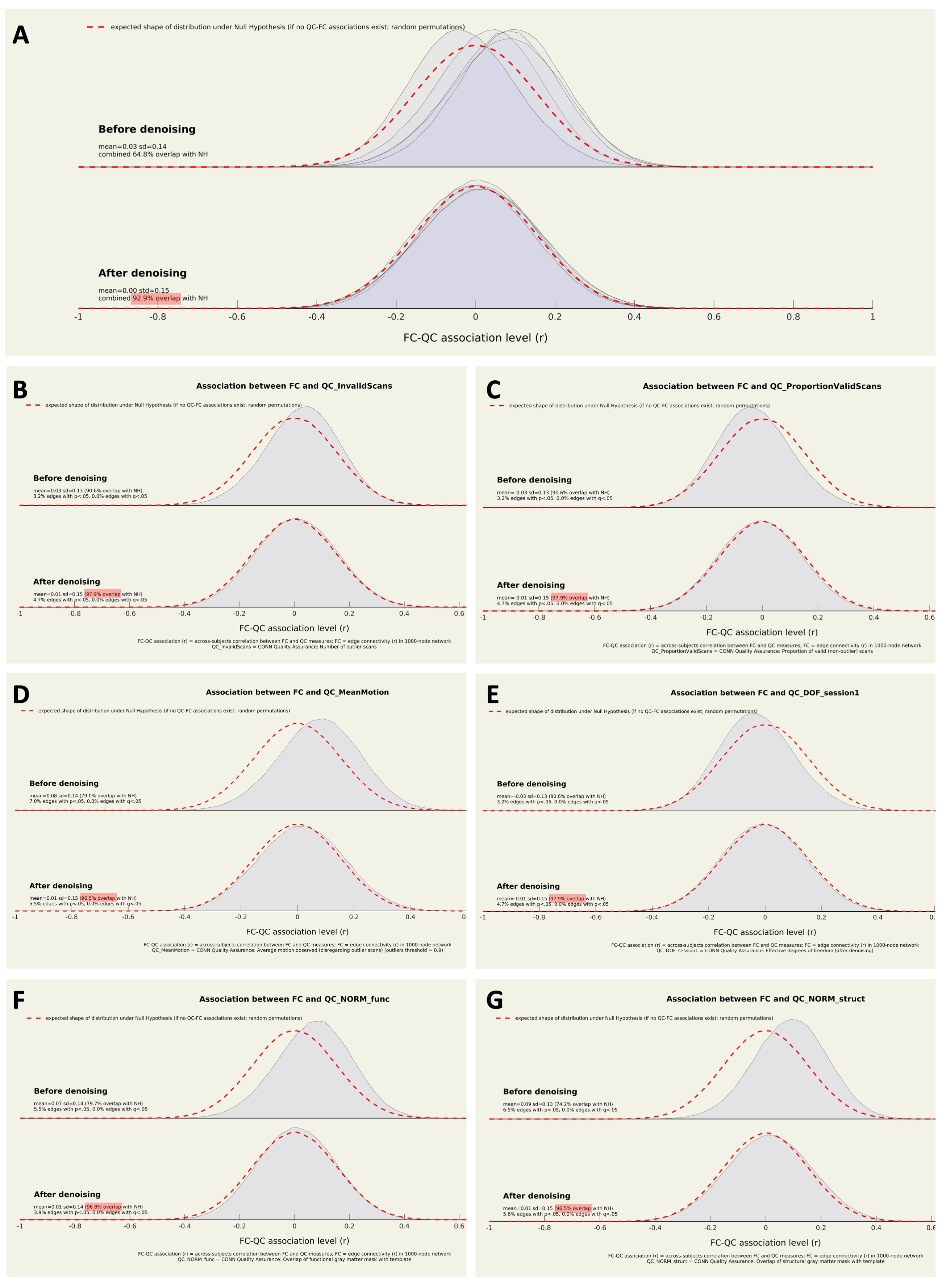

*Note: Detailed description of QC parameters see Section 2.1; A) Distribution of QC-FC associations (QC-FC) over all denoising parameters; Association between FC and B) Invalid Scans; C) Proportion of Valid Scans; D) Mean Motion; E) DOF (= effective degrees of freedom of the BOLD timeseries after denoising); F) normalization (NORM_func); G) anatomical normalization (NORM_anat); NORM_func and NORM_anat measured the similarity between the gray matter mask in the normalized data and in a reference MNI atlas. QC, Quality control; FC, Functional Connectivity*

Figure S5: Results of Ridge regression

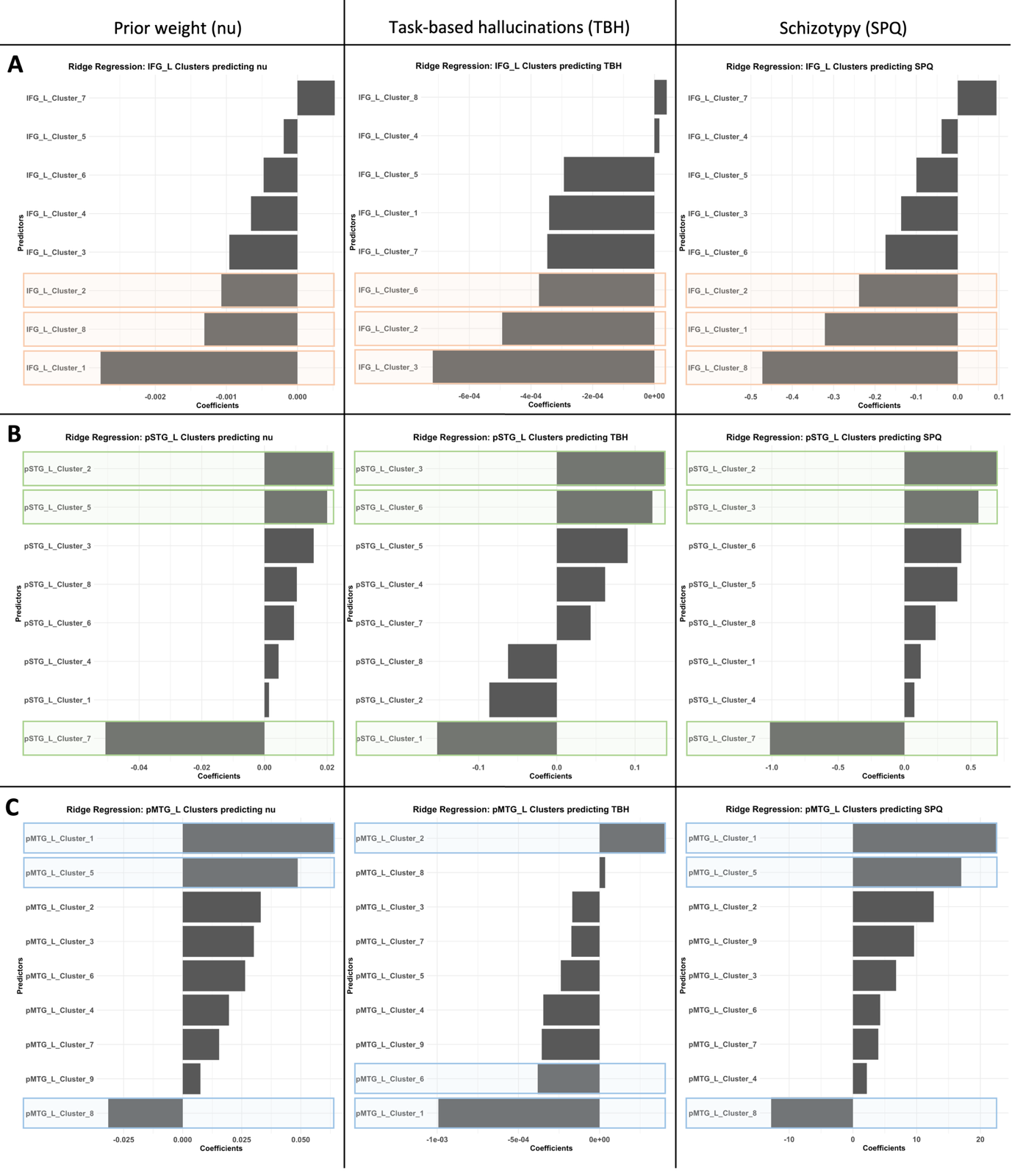

*Note: Identifying the three most significant predictors with a ridge regression for prior weight (nu) in column one, task-based hallucinations (TBH) in column two, and schizotypy (SPQ) in column three. A) shows the results for the significant Clusters of the left IFG (orange), B) for the left pSTG (green), and C) for the left pMTG (blue).*

*nu, Prior weight of our computational model; SPQ, schizotypy personality questionnaire; IFG, Inferior Frontal Gyrus; pSTG, posterior Superior Temporal Gyrus; pMTG, posterior Middle Temporal Gyrus*

Figure S6: Bayesian regression model – quality checks

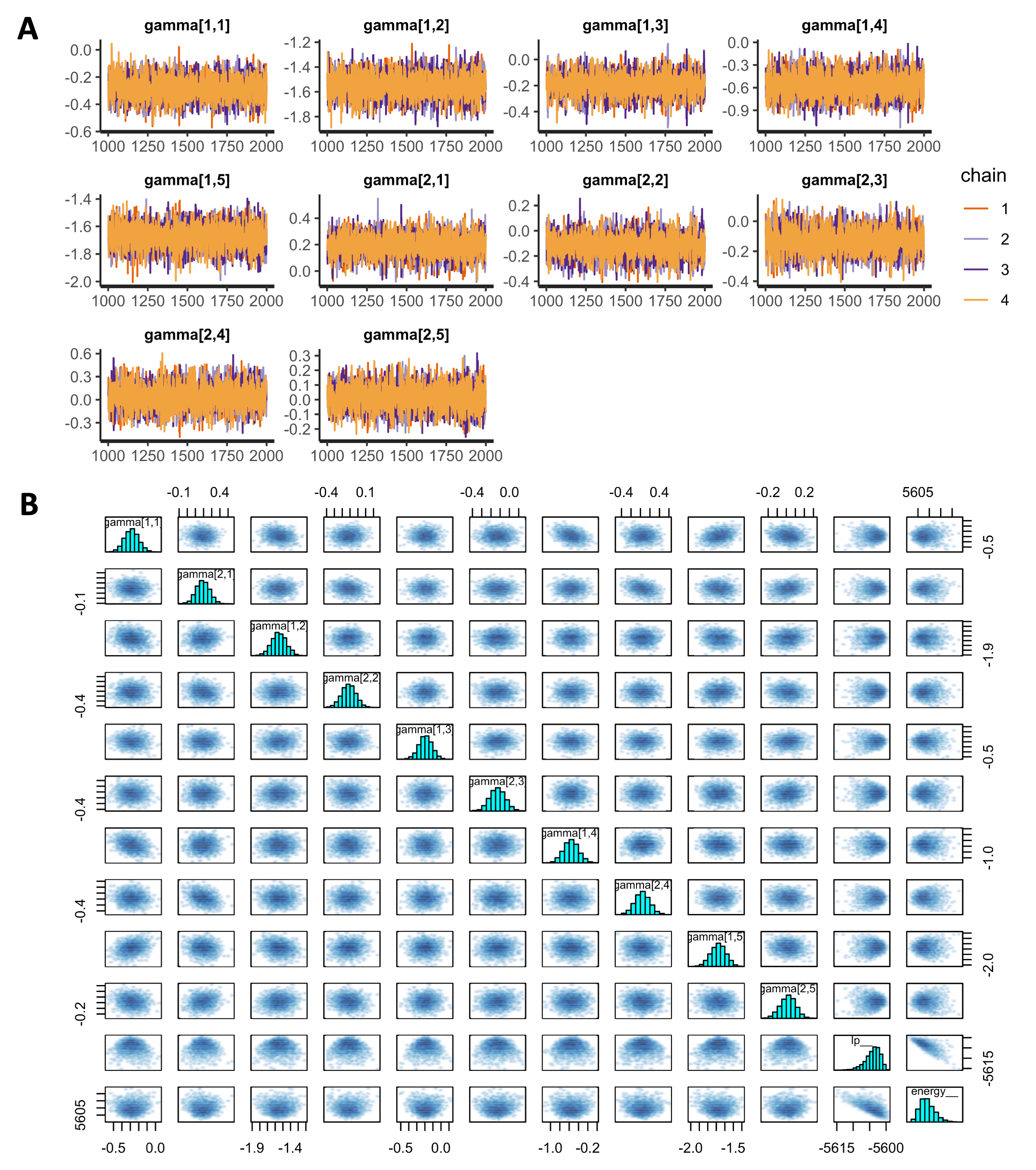

*Note: A) Trace plots for direct-effect model with individual effects for 53 participants for each population gamma showing successful convergence of four Markov Chains, each with 2000 iterations (warmup=1000; thin=1; post-warmup draws per chain=1000, total post-warmup draws=4000 for each gamma); B)* *Pairwise scatterplots and marginal posterior distributions of population gammas. Diagonal histograms indicate normal distributions for all gammas. Scatterplot indicate little to no correlations between gammas.*

Figure S7: Influence of increasing schizotypy on prior weight

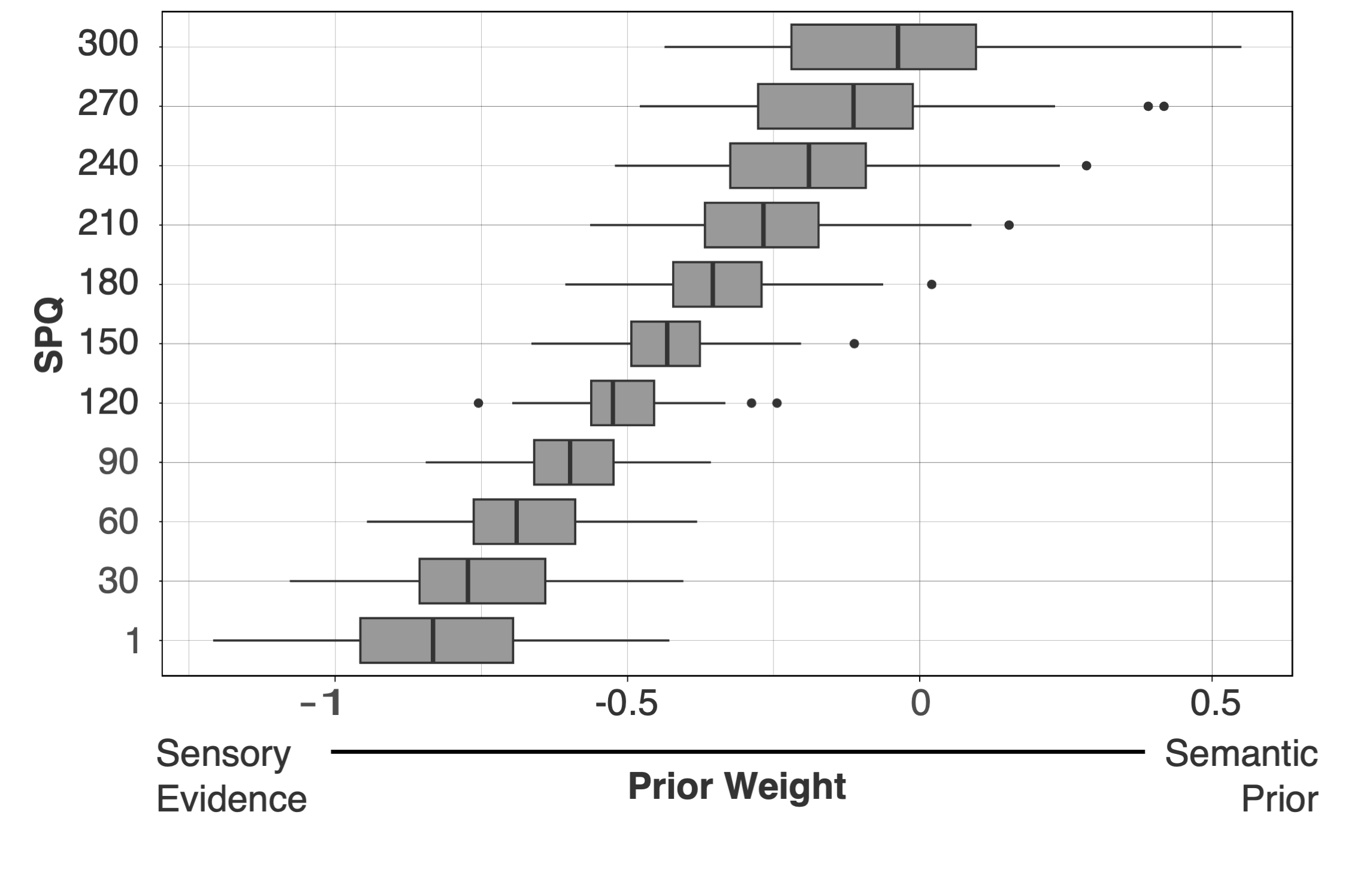

*Note: Posterior predictive simulation of the relative prior weight 𝜈i for an average task trial when systematically increasing SPQ. The simulation illustrates the overweighting of prior beliefs relative to sensory evidence with increasing schizotypy.*

### 4 **Tables**

Table S1: Demographic data, clinical scores, and behavioural data.

|  | **Female (n = 26)** | **Male (n = 27)** | **P-value**^a^ | **Test Statistic** |
| --- | --- | --- | --- | --- |
| Age | 23.31 (3.54) | 23.93 (4.21) | 0.7266 | W=331^a^ |
| Total Score SPQ | 94.31 (45.76) | 82.85 (53.78) | 0.2777 | W=412.5^a^ |
| Prior weight (nu) | -0.48 (0.26) | -0.54 (0.23) | 0.3446 | t(50.20) = 0.95^b^ |
| Task-based hallucinations | 0.15 (0.07) | 0.14 (0.06) | 0.5564 | t(49.98) = 0.59^b^ |
| *Note: Values are mean (SD)*  *SPQ, Schizotypal Personality Questionnaire; AQ, Autism Spectrum Quotient* | | | | |

^a^ *Wilcoxon rank sum test with continuity correction, ^b^Welch Two Sample t-test*

Table S2: Composition of significant clusters correlating with the left IFG

| Cluster | % of voxels in the cluster | % of voxels of the region covered | Region | Center coordinates |
| --- | --- | --- | --- | --- |
| 1/8 | 14 | 93 | IFG oper l (Inferior Frontal Gyrus, pars opercularis Left) | -50,+14,+16 |
| 1/8 | 10 | 81 | IFG tri l (Inferior Frontal Gyrus, pars triangularis Left) | -50,+28,+8 |
| 1/8 | 15 | 45 | FOrb l (Frontal Orbital Cortex Left) | -38,+24,-14 |
| 1/8 | 3 | 38 | FO l (Frontal Operculum Cortex Left) | -44,+20,+0 |
| 1/8 | 20 | 35 | MidFG l (Middle Frontal Gyrus Left) | -42,+16,+42 |
| 1/8 | 12 | 9 | FP l (Frontal Pole Left) | -44,+44,-4 |
| 1/8 | 4 | 9 | TP l (Temporal Pole Left) | -44,+18,-20 |
| 1/8 | 7 | 8 | PreCG l (Precentral Gyrus Left) | -48,+4,+26 |
| 1/8 | 1 | 4 | IC l (Insular Cortex Left) | -34,+22,-4 |
| 1/8 | 15 | 0 | not-labeled | -42,+24,+8 |
| 2/8 | 30 | 65 | pMTG l (Middle Temporal Gyrus, posterior division Left) | -60,-32,-8 |
| 2/8 | 14 | 44 | AG l (Angular Gyrus Left) | -52,-52,+30 |
| 2/8 | 14 | 40 | pSMG l (Supramarginal Gyrus, posterior division Left) | -54,-48,+26 |
| 2/8 | 5 | 38 | pSTG l (Superior Temporal Gyrus, posterior division Left) | -58,-32,+2 |
| 2/8 | 9 | 30 | toMTG l (Middle Temporal Gyrus, temporooccipital part Left) | -60,-48,+2 |
| 2/8 | 4 | 2 | sLOC l (Lateral Occipital Cortex, superior division Left) | -40,-62,+52 |
| 2/8 | 1 | 2 | SPL l (Superior Parietal Lobule Left) | -38,-54,+54 |
| 2/8 | 23 | 0 | not-labeled | -48,-44,+14 |
| 2/8 | 0 | 0 | pITG l (Inferior Temporal Gyrus, posterior division Left) | -64,-36,-20 |
| 3/8 | 23 | 24 | SFG l (Superior Frontal Gyrus Left) | -8,+34,+50 |
| 3/8 | 16 | 17 | SFG r (Superior Frontal Gyrus Right) | +6,+34,+50 |
| 3/8 | 18 | 8 | FP l (Frontal Pole Left) | -12,+50,+38 |
| 3/8 | 3 | 6 | PaCiG l (Paracingulate Gyrus Left) | -4,+36,+36 |
| 3/8 | 14 | 5 | FP r (Frontal Pole Right) | +12,+50,+38 |
| 3/8 | 2 | 3 | PaCiG r (Paracingulate Gyrus Right) | +4,+38,+36 |
| 3/8 | 25 | 0 | not-labeled | +0,+40,+46 |
| 4/8 | 20 | 73 | IFG tri r (Inferior Frontal Gyrus, pars triangularis Right) | +52,+26,+8 |
| 4/8 | 17 | 52 | IFG oper r (Inferior Frontal Gyrus, pars opercularis Right) | +54,+18,+12 |
| 4/8 | 19 | 27 | FOrb r (Frontal Orbital Cortex Right) | +42,+26,-12 |
| 4/8 | 3 | 17 | FO r (Frontal Operculum Cortex Right) | +46,+20,+0 |
| 4/8 | 18 | 5 | FP r (Frontal Pole Right) | +46,+40,-8 |
| 4/8 | 6 | 5 | TP r (Temporal Pole Right) | +48,+18,-16 |
| 4/8 | 17 | 0 | not-labeled | +50,+26,+2 |
| 4/8 | 0 | 0 | MidFG r (Middle Frontal Gyrus Right) | +52,+26,+24 |
| 5/8 | 55 | 32 | Cereb2 r (Cerebelum Crus2 Right) | +20,-80,-38 |
| 5/8 | 11 | 25 | Cereb7 r (Cerebelum 7b Right) | +24,-76,-48 |
| 5/8 | 26 | 13 | Cereb1 r (Cerebelum Crus1 Right) | +22,-74,-30 |
| 5/8 | 1 | 8 | Ver7 (Vermis 7) | +4,-78,-24 |
| 5/8 | 1 | 1 | Cereb2 l (Cerebelum Crus2 Left) | +0,-82,-28 |
| 5/8 | 2 | 1 | Cereb8 r (Cerebelum 8 Right) | +24,-72,-48 |
| 5/8 | 4 | 0 | not-labeled | +24,-80,-52 |
| 6/8 | 21 | 38 | pSTG r (Superior Temporal Gyrus, posterior division Right) | +60,-26,+0 |
| 6/8 | 49 | 28 | pMTG r (Middle Temporal Gyrus, posterior division Right) | +60,-28,-4 |
| 6/8 | 15 | 10 | toMTG r (Middle Temporal Gyrus, temporooccipital part Right) | +60,-40,+2 |
| 6/8 | 2 | 1 | pSMG r (Supramarginal Gyrus, posterior division Right) | +56,-40,+6 |
| 6/8 | 14 | 0 | not-labeled | +58,-30,+4 |
| 7/8 | 40 | 30 | aMTG l (Middle Temporal Gyrus, anterior division Left) | -56,+0,-26 |
| 7/8 | 45 | 6 | TP l (Temporal Pole Left) | -52,+4,-30 |
| 7/8 | 4 | 5 | aSTG l (Superior Temporal Gyrus, anterior division Left) | -52,+2,-16 |
| 7/8 | 12 | 0 | not-labeled | -48,+2,-30 |
| 7/8 | 0 | 0 | aITG l (Inferior Temporal Gyrus, anterior division Left) | -46,+0,-34 |
| 8/8 | 63 | 8 | Cereb2 l (Cerebelum Crus2 Left) | -18,-82,-34 |
| 8/8 | 8 | 4 | Cereb7 l (Cerebelum 7b Left) | -22,-74,-46 |
| 8/8 | 27 | 3 | Cereb1 l (Cerebelum Crus1 Left) | -22,-80,-28 |
| 8/8 | 3 | 0 | not-labeled | -20,-78,-50 |

Table S3: Composition of significant clusters correlation with the left pSTG

| Cluster | % of voxels in the cluster | % of voxels of the region covered | Region | Center coordinates |
| --- | --- | --- | --- | --- |
| 1/8 | 4 | 99 | aMTG l (Middle Temporal Gyrus, anterior division Left) | -58,-4,-22 |
| 1/8 | 11 | 82 | pMTG l (Middle Temporal Gyrus, posterior division Left) | -60,-28,-10 |
| 1/8 | 2 | 60 | aITG l (Inferior Temporal Gyrus, anterior division Left) | -48,-4,-38 |
| 1/8 | 3 | 54 | IFG tri l (Inferior Frontal Gyrus, pars triangularis Left) | -52,+26,+6 |
| 1/8 | 5 | 52 | pSMG l (Supramarginal Gyrus, posterior division Left) | -58,-46,+24 |
| 1/8 | 4 | 52 | toMTG l (Middle Temporal Gyrus, temporooccipital part Left) | -60,-50,+4 |
| 1/8 | 2 | 50 | pSTG l (Superior Temporal Gyrus, posterior division Left) | -60,-30,+2 |
| 1/8 | 8 | 48 | FOrb l (Frontal Orbital Cortex Left) | -38,+24,-14 |
| 1/8 | 3 | 48 | IFG oper l (Inferior Frontal Gyrus, pars opercularis Left) | -50,+16,+16 |
| 1/8 | 9 | 42 | TP l (Temporal Pole Left) | -44,+12,-28 |
| 1/8 | 1 | 40 | aSTG l (Superior Temporal Gyrus, anterior division Left) | -54,-4,-12 |
| 1/8 | 8 | 28 | MidFG l (Middle Frontal Gyrus Left) | -40,+14,+46 |
| 1/8 | 1 | 27 | FO l (Frontal Operculum Cortex Left) | -44,+22,+0 |
| 1/8 | 8 | 16 | sLOC l (Lateral Occipital Cortex, superior division Left) | -46,-64,+36 |
| 1/8 | 1 | 7 | IC l (Insular Cortex Left) | -34,+16,-10 |
| 1/8 | 3 | 5 | FP l (Frontal Pole Left) | -44,+42,-6 |
| 1/8 | 0 | 5 | pITG l (Inferior Temporal Gyrus, posterior division Left) | -54,-14,-32 |
| 1/8 | 0 | 3 | PO l (Parietal Operculum Cortex Left) | -54,-40,+24 |
| 1/8 | 0 | 2 | aTFusC l (Temporal Fusiform Cortex, anterior division Left) | -38,-4,-46 |
| 1/8 | 18 | 1 | not-labeled | -48,-22,+0 |
| 1/8 | 0 | 1 | PT l (Planum Temporale Left) | -52,-42,+18 |
| 1/8 | 0 | 0 | iLOC l (Lateral Occipital Cortex, inferior division Left) | -48,-64,+14 |
| 1/8 | 0 | 0 | SPL l (Superior Parietal Lobule Left) | -36,-58,+48 |
| 2/8 | 18 | 40 | SFG l (Superior Frontal Gyrus Left) | -8,+32,+52 |
| 2/8 | 9 | 22 | SFG r (Superior Frontal Gyrus Right) | +8,+34,+52 |
| 2/8 | 23 | 21 | FP l (Frontal Pole Left) | -18,+50,+32 |
| 2/8 | 4 | 19 | PaCiG l (Paracingulate Gyrus Left) | -6,+44,+24 |
| 2/8 | 21 | 16 | FP r (Frontal Pole Right) | +16,+54,+30 |
| 2/8 | 2 | 9 | PaCiG r (Paracingulate Gyrus Right) | +4,+44,+26 |
| 2/8 | 22 | 0 | not-labeled | +0,+42,+40 |
| 2/8 | 0 | 0 | SMA L(Juxtapositional Lobule Cortex -formerly Supplementary Motor Cortex- Left) | -6,+8,+62 |
| 3/8 | 9 | 91 | aMTG r (Middle Temporal Gyrus, anterior division Right) | +58,-2,-26 |
| 3/8 | 22 | 67 | pMTG r (Middle Temporal Gyrus, posterior division Right) | +60,-22,-10 |
| 3/8 | 5 | 63 | aITG r (Inferior Temporal Gyrus, anterior division Right) | +48,-4,-38 |
| 3/8 | 3 | 35 | pSTG r (Superior Temporal Gyrus, posterior division Right) | +60,-24,-2 |
| 3/8 | 15 | 27 | TP r (Temporal Pole Right) | +46,+12,-28 |
| 3/8 | 2 | 24 | aSTG r (Superior Temporal Gyrus, anterior division Right) | +52,+0,-18 |
| 3/8 | 8 | 24 | FOrb r (Frontal Orbital Cortex Right) | +40,+24,-14 |
| 3/8 | 8 | 23 | AG r (Angular Gyrus Right) | +56,-52,+20 |
| 3/8 | 5 | 18 | toMTG r (Middle Temporal Gyrus, temporooccipital part Right) | +58,-42,+6 |
| 3/8 | 2 | 8 | pITG r (Inferior Temporal Gyrus, posterior division Right) | +50,-10,-36 |
| 3/8 | 2 | 6 | pSMG r (Supramarginal Gyrus, posterior division Right) | +50,-42,+12 |
| 3/8 | 2 | 1 | FP r (Frontal Pole Right) | +46,+36,-14 |
| 3/8 | 0 | 1 | IFG tri r (Inferior Frontal Gyrus, pars triangularis Right) | +52,+28,-6 |
| 3/8 | 0 | 1 | PP r (Planum Polare Right) | +48,+2,-16 |
| 3/8 | 16 | 0 | not-labeled | +50,-12,-12 |
| 3/8 | 1 | 0 | sLOC r (Lateral Occipital Cortex, superior division Right) | +58,-60,+24 |
| 3/8 | 0 | 0 | pTFusC r (Temporal Fusiform Cortex, posterior division Right) | +44,-10,-38 |
| 4/8 | 52 | 30 | Cereb2 r (Cerebelum Crus2 Right) | +20,-82,-36 |
| 4/8 | 40 | 19 | Cereb1 r (Cerebelum Crus1 Right) | +24,-78,-30 |
| 4/8 | 1 | 1 | Cereb6 r (Cerebelum 6 Right) | +20,-72,-26 |
| 4/8 | 0 | 1 | Cereb7 r (Cerebelum 7b Right) | +16,-76,-42 |
| 4/8 | 0 | 1 | Ver7 (Vermis 7) | +6,-76,-26 |
| 4/8 | 6 | 0 | not-labeled | +22,-90,-40 |
| 4/8 | 0 | 0 | Cereb8 r (Cerebelum 8 Right) | +16,-70,-36 |
| 5/8 | 88 | 10 | Precuneous (Precuneous Cortex) | -2,-56,+38 |
| 5/8 | 9 | 2 | PC (Cingulate Gyrus, posterior division) | -6,-48,+34 |
| 5/8 | 3 | 0 | not-labeled | -12,-58,+36 |
| 6/8 | 74 | 5 | FP l (Frontal Pole Left) | -10,+62,-14 |
| 6/8 | 3 | 1 | MedFC (Frontal Medial Cortex) | -4,+54,-16 |
| 6/8 | 15 | 0 | not-labeled | -8,+56,-14 |
| 6/8 | 8 | 0 | FP r (Frontal Pole Right) | +4,+60,-14 |
| 7/8 | 100 | 0 | not-labeled; natbrainlab: Corpus Callosum left | -18,+14,+24 |
| 8/8 | 73 | 10 | Cereb2 l (Cerebelum Crus2 Left) | -26,-80,-36 |
| 8/8 | 27 | 3 | Cereb1 l (Cerebelum Crus1 Left) | -26,-76,-30 |

Table S4: Composition of significant clusters correlation with the left pMTG

| Cluster | % of voxels in the cluster | % of voxels of the region covered | Region | Center coordinates |
| --- | --- | --- | --- | --- |
| 1/9 | 3 | 95 | aMTG l (Middle Temporal Gyrus, anterior division Left) | -58,-4,-22 |
| 1/9 | 5 | 75 | AG l (Angular Gyrus Left) | -50,-56,+26 |
| 1/9 | 1 | 45 | aITG l (Inferior Temporal Gyrus, anterior division Left) | -48,-6,-38 |
| 1/9 | 1 | 41 | pSTG l (Superior Temporal Gyrus, posterior division Left) | -60,-28,+0 |
| 1/9 | 8 | 40 | MidFG l (Middle Frontal Gyrus Left) | -40,+16,+44 |
| 1/9 | 2 | 38 | IFG oper l (Inferior Frontal Gyrus, pars opercularis Left) | -52,+18,+18 |
| 1/9 | 7 | 36 | SFG l (Superior Frontal Gyrus Left) | -10,+34,+48 |
| 1/9 | 6 | 35 | TP l (Temporal Pole Left) | -42,+10,-30 |
| 1/9 | 4 | 35 | FOrb l (Frontal Orbital Cortex Left) | -38,+22,-16 |
| 1/9 | 1 | 34 | IFG tri l (Inferior Frontal Gyrus, pars triangularis Left) | -52,+24,+6 |
| 1/9 | 0 | 25 | aSTG l (Superior Temporal Gyrus, anterior division Left) | -54,-4,-14 |
| 1/9 | 10 | 23 | FP l (Frontal Pole Left) | -14,+54,+26 |
| 1/9 | 2 | 23 | PaCiG l (Paracingulate Gyrus Left) | -6,+46,+18 |
| 1/9 | 1 | 21 | toMTG l (Middle Temporal Gyrus, temporooccipital part Left) | -62,-48,+2 |
| 1/9 | 6 | 19 | sLOC l (Lateral Occipital Cortex, superior division Left) | -44,-66,+40 |
| 1/9 | 3 | 19 | SFG r (Superior Frontal Gyrus Right) | +6,+40,+44 |
| 1/9 | 1 | 17 | pSMG l (Supramarginal Gyrus, posterior division Left) | -56,-48,+18 |
| 1/9 | 8 | 15 | FP r (Frontal Pole Right) | +14,+56,+28 |
| 1/9 | 1 | 11 | PaCiG r (Paracingulate Gyrus Right) | +6,+48,+20 |
| 1/9 | 0 | 7 | FO l (Frontal Operculum Cortex Left) | -46,+22,+0 |
| 1/9 | 0 | 6 | pITG l (Inferior Temporal Gyrus, posterior division Left) | -62,-32,-20 |
| 1/9 | 0 | 6 | aTFusC l (Temporal Fusiform Cortex, anterior division Left) | -36,+0,-40 |
| 1/9 | 0 | 4 | MedFC (Frontal Medial Cortex) | -2,+54,-14 |
| 1/9 | 0 | 2 | IC l (Insular Cortex Left) | -32,+12,-16 |
| 1/9 | 19 | 1 | not-labeled | -26,+20,+32 |
| 1/9 | 0 | 1 | AC (Cingulate Gyrus, anterior division) | -8,+42,+8 |
| 1/9 | 0 | 0 | PreCG l (Precentral Gyrus Left) | -42,+4,+40 |
| 1/9 | 0 | 0 | SPL l (Superior Parietal Lobule Left) | -36,-58,+48 |
| 1/9 | 0 | 0 | MidFG r (Middle Frontal Gyrus Right) | +26,+32,+48 |
| 2/9 | 10 | 86 | aMTG r (Middle Temporal Gyrus, anterior division Right) | +56,-2,-26 |
| 2/9 | 32 | 84 | pMTG r (Middle Temporal Gyrus, posterior division Right) | +62,-22,-12 |
| 2/9 | 5 | 53 | aITG r (Inferior Temporal Gyrus, anterior division Right) | +48,-4,-38 |
| 2/9 | 5 | 39 | pSTG r (Superior Temporal Gyrus, posterior division Right) | +60,-26,+0 |
| 2/9 | 16 | 24 | TP r (Temporal Pole Right) | +42,+12,-30 |
| 2/9 | 2 | 21 | aSTG r (Superior Temporal Gyrus, anterior division Right) | +52,+0,-18 |
| 2/9 | 7 | 19 | FOrb r (Frontal Orbital Cortex Right) | +40,+26,-14 |
| 2/9 | 3 | 13 | pITG r (Inferior Temporal Gyrus, posterior division Right) | +50,-12,-34 |
| 2/9 | 2 | 7 | toMTG r (Middle Temporal Gyrus, temporooccipital part Right) | +62,-40,+0 |
| 2/9 | 0 | 1 | pTFusC r (Temporal Fusiform Cortex, posterior division Right) | +42,-12,-38 |
| 2/9 | 18 | 0 | not-labeled | +50,-12,-20 |
| 2/9 | 1 | 0 | FP r (Frontal Pole Right) | +38,+34,-14 |
| 2/9 | 0 | 0 | IC r (Insular Cortex Right) | +28,+14,-14 |
| 3/9 | 32 | 22 | PC (Cingulate Gyrus, posterior division) | -2,-46,+32 |
| 3/9 | 56 | 17 | Precuneous (Precuneous Cortex) | -2,-58,+36 |
| 3/9 | 13 | 0 | not-labeled | -8,-46,+28 |
| 4/9 | 57 | 42 | Cereb2 r (Cerebelum Crus2 Right) | +20,-80,-36 |
| 4/9 | 37 | 23 | Cereb1 r (Cerebelum Crus1 Right) | +28,-76,-30 |
| 4/9 | 1 | 2 | Cereb7 r (Cerebelum 7b Right) | +18,-76,-44 |
| 4/9 | 1 | 1 | Cereb2 l (Cerebelum Crus2 Left) | +0,-84,-28 |
| 4/9 | 5 | 0 | not-labeled | +12,-90,-38 |
| 5/9 | 65 | 20 | AG r (Angular Gyrus Right) | +50,-52,+24 |
| 5/9 | 28 | 3 | sLOC r (Lateral Occipital Cortex, superior division Right) | +54,-62,+34 |
| 5/9 | 7 | 0 | not-labeled | +42,-48,+24 |
| 6/9 | 68 | 13 | Cereb2 l (Cerebelum Crus2 Left) | -26,-80,-36 |
| 6/9 | 32 | 5 | Cereb1 l (Cerebelum Crus1 Left) | -28,-76,-30 |
| 7/9 | 75 | 3 | FP l (Frontal Pole Left) | -38,+46,-4 |
| 7/9 | 25 | 0 | not-labeled | -32,+46,-4 |
| 8/9 | 84 | 2 | FP r (Frontal Pole Right) | +42,+42,+24 |
| 8/9 | 12 | 1 | MidFG r (Middle Frontal Gyrus Right) | +38,+34,+28 |
| 8/9 | 4 | 0 | not-labeled | +38,+34,+24 |
| 9/9 | 91 | 0 | not-labeled | -30,-72,+2 |
| 9/9 | 5 | 0 | iLOC l (Lateral Occipital Cortex, inferior division Left) | -38,-78,+8 |
| 9/9 | 2 | 0 | OFusG l (Occipital Fusiform Gyrus Left) | -28,-70,-4 |
| 9/9 | 2 | 0 | sLOC l (Lateral Occipital Cortex, superior division Left) | -38,-78,+14 |
